# Ropinirole hydrochloride for amyotrophic lateral sclerosis: A single-center, randomized feasibility, double-blind, placebo-controlled trial

**DOI:** 10.1101/2021.12.05.21267266

**Authors:** Satoru Morimoto, Shinichi Takahashi, Daisuke Ito, Yugaku Daté, Kensuke Okada, Chai Muh Chyi, Ayumi Nishiyama, Naoki Suzuki, Koki Fujimori, Masaki Takao, Miwa Hirai, Yasuaki Kabe, Makoto Suematsu, Masahiro Jinzaki, Masashi Aoki, Yuto Fujiki, Yasunori Sato, Norihiro Suzuki, Jin Nakahara, Hideyuki Okano

## Abstract

**Background:** We previously used an induced pluripotent stem cell-based drug repurposing approach to demonstrate that ropinirole hydrochloride (ropinirole) attenuated amyotrophic lateral sclerosis (ALS)-specific pathological phenotypes. Here, we assessed the safety and feasibility of ropinirole in ALS patients to verify its efficacy.

**Methods:** We conducted a randomized feasibility trial of ALS. Twenty participants with ALSFRS-R scores greater than 2 points were randomly assigned using dynamic allocation to receive ropinirole or placebo for 24 weeks in the double-blind period. Upon completion, participants could choose to participate in the following 24-week open-label active extension period. The primary outcomes were safety and tolerability. The secondary outcomes for the feasibility trial objective were the change in the ALS Functional Rating Scale-Revised (ALSFRS-R) score, composite functional endpoint, combined assessment of function and survival, event-free survival, and time to ≤50% forced vital capacity (blinded outcome assessment). This study is registered with the UMIN Clinical Trials Registry, UMIN000034954.

**Findings:** Twenty-one participants were randomized into two groups (ropinirole group; n=14) and received ropinirole (n=13) or placebo (n=7) and the data of all participants were analysed using mixed-effects models for repeated measures together. Overall, the incidences of adverse events, most of which had been reported previously, were similar within both groups. Notably, the incidence of gastrointestinal disorders (mainly, temporary mild nausea and diarrhoea) was high at 76·9% in the ropinirole group (14·3% in the placebo group). Regarding the feasibility of verifying efficacy, there were no significant differences in the ALSFRS-R score and combined assessment of function and survival scores during the double-blind period for 6 months, while the participants in the ropinirole group had lived an additional 27·9 weeks without disease progression events compared with the placebo group (log-rank test, 95% confidence interval, 4·3–37·4) at 12 months (secondary outcome).

**Interpretation:** Ropinirole is safe and tolerable for patients with ALS and this trial indicates feasibility for a subsequent large-scale trial.

**Funding:** This study was funded by The Japan Agency for Medical Research and Development and K Pharma Inc.

## 1 INTRODUCTION

Amyotrophic lateral sclerosis (ALS) is a neurodegenerative disease. The median survival time of patients with ALS is approximately 2 years, and muscle weakness eventually causes respiratory failure and death.^1^ Currently, there are two Food and Drug Administration-approved drugs for the treatment of ALS: riluzole and edaravone. Riluzole extends survival by 2–3 months with no reported benefits to muscle strength, whereas edaravone improves short-term functional outcomes but has no statistically significant effect on survival.^2^ Therefore, the need for effective ALS therapies remains unmet.

Drug development for diseases of the nervous system has the second-lowest success rate at the pivotal phase compared with other therapeutic areas.^3^ In particular, neurodegenerative diseases are heterogeneous and the majority of them is sporadic, limiting the translational potential of preclinical animal models. By exploiting induced pluripotent stem cell (iPSC)-derived motor neurons (MNs) generated from patients with ALS, we sought to enable large-scale drug screening and overcome the limitations of preclinical animal models for the development of drugs to treat highly heterogeneous neurodegenerative diseases.^4^

Ropinirole hydrochloride (ropinirole) was identified as a treatment candidate for ALS from 1232 FDA-approved drugs in a drug screening analysis conducted at Keio University. The analysis examined 3 *FUS*- and 2 *TDP-43* (*TARDBP*)-ALS patient iPSC-derived lower MNs (LMNs) for the suppression of ALS-related phenotypes *in vitro*, such as MN death and damage, neurite regression, mislocalization of FUS and TDP-43, and stress granule formation. Furthermore, we established iPSCs derived from 32 sporadic ALS patients and induced LMNs. Among 32 LMNs, 22 of 24 LMNs with cell damage and neurite regression demonstrated increased apoptosis.

Finally, we confirmed that ropinirole hydrochloride improved multiple phenotypes of ALS in 16 of 22 LMNs (72·73%).^5^

Ropinirole is a commonly used drug for Parkinson’s disease (PD) that can permeate the blood-brain barrier. Of note, our *in vitro* model suggested that ropinirole was more potent than other ALS drugs and investigational drugs, including riluzole, edaravone, and ceftriaxone.^5^ Interestingly, other dopamine D2 receptor agonists such as bromocriptine and pramipexole

(dexpramipexole), have been reported to be candidates for therapeutics for ALS, ^6,7^ but these drugs failed to show efficacy in clinical trials had.^8-11^

Based on the safety profile of ropinirole for PD, we hypothesized that ropinirole would be well tolerated in patients with ALS. Here, we present the results of the Ropinirole Hydrochloride Remedy for Amyotrophic Lateral Sclerosis (ROPALS) single-centre, randomized, placebo-controlled trial, which evaluated the safety and efficacy of ropinirole with the following parameters: ALS Functional Rating Scale-Revised (ALSFRS-R) score,^12^ composite functional endpoint, event-free survival, and time to ≤50% forced vital capacity (FVC), of the ropinirole hydrochloride extended-release tablet (Requip CR) in participants with ALS.^13,14^

## 2 METHODS

### 2.1 Study design and patients

The ROPALS trial was an investigator-led, double-blind, placebo-controlled, randomized phase 1/2a trial conducted at a single site (Keio University Hospital) in Japan beginning in Dec 2018 (figure S1). This trial was approved by the Institutional Review Board of Keio University Hospital (No. D18-01). Participating investigators are listed in Section S1. Participants were followed up for 64 weeks, which comprised a run-in period of 12 weeks followed by a 48-week intervention period and a 4-week follow-up period (figure S1).

We conducted this trial in accordance with the Good Clinical Practice guidelines of the International Council for Harmonisation, and the trial protocol was approved by our institutional review board. The trial protocol has been published,^15^ and the first and last versions and history of changes in the protocol and statistical analysis plan are available as supplementary files. An independent data monitoring committee was set up for this clinical trial. Written informed consent was provided by participants or their legal representatives. GlaxoSmithKline K.K. (GSK) provided ropinirole extended-release tablets and placebo. Data were collected and analysed by the investigators and an independent company (DOT WORLD Co., Ltd., Tokyo, Japan).

This trial included patients with ALS who satisfied all the inclusion criteria (patients with a change in the ALSFRS-R score within the range of −2 to −5 points during the 12-week run-in period at official registration) and did not meet any of the exclusion criteria (patients with a family history or prior diagnosis of *superoxide dismutase (SOD)-1* mutation). Furthermore, concomitant use of riluzole was allowed during the period from obtaining informed consent to the end of the study or to the time of discontinuation. Participants who were not receiving riluzole before providing informed consent were not allowed to start treatment with riluzole after providing informed consent, and the use of edaravone was prohibited. Full details of the inclusion and exclusion criteria are provided in the protocol.

### 2.2 SAMPLE SIZE

With 20 patients (15 and 5 in the treatment and control groups, respectively), a clinically important adverse event (AE) with a 10% incidence rate in the treatment group can be detected with 80% probability. For the efficacy evaluation, the mean change in ALSFRS-R score from baseline over 24 weeks in the treatment group was assumed to be −5·5 (standard deviation = 6·0). Under this assumption, with 80% probability, the sample mean change in the treatment group in this trial was greater than the threshold of −6·8, calculated from the placebo group in previous confirmatory trials (n = 99 and n = 66) of ALS patients.^16,17^

### 2.3 Randomization and masking

After screening, participants were randomly assigned to the active drug or placebo group at a 3:1 ratio by DOT WORLD Co. Ltd. through a computer-generated process and web response system with dynamic allocation and minimisation for the number of months after onset (<30 months vs. ≥30 months), age (<60 years vs. ≥60 years), and total ALSFRS-R score (≤36 vs. ≥37). The study drugs were assigned and labelled with random numbers according to the randomization table created by DOT WORLD Co. Ltd. personnel who were not involved in conducting the trial or performing the analysis. Patients, investigators, and study staff were not able to access the table and were masked to treatment group assignments.

### 2.4 Procedures

Study treatment was started at an initial dose of 2 mg per day, followed by increases in the dose once weekly to a maximum of 16 mg per day. After double-blind and open-label extension periods, the dose of the study drug was tapered in accordance with the protocol. Patients were assessed by neurologists for functional grade and neurological and laboratory (blood and urine) findings during the screening period at 4 (only ALSFRS-R and the amount of physical activity), 8 (only ALSFRS-R and the amount of physical activity), and 12 weeks after interim registration; before the start of the first dose of study treatment; at weeks 5, 9, 13, 17, 21, 24, 27, 31, 35, 39, 43, 47, and 50 after the start of study treatment; during the follow-up period; and at the time of discontinuation. The amount of physical activity (METs: metabolic equivalents) was measured every 10 seconds by Active style Pro HJA-750C with a high precision 3D accelerometer (OMRON, Kyoto, Japan) and evaluated as the 4-week mean value. In addition, cerebrospinal fluid (CSF) tests were performed using lumbar puncture before the start of the first dose of the study treatment, before study drug administration at weeks 24 and 50 after the start of study treatment, and at the time of discontinuation. A detailed schedule of the assessments is provided in the protocol. Data were collected with an electronic case-report form using Viedoc (Viedoc Japan K.K., Tokyo, Japan).

### 2.5 Outcomes

In the randomized study, the primary outcomes were AEs, laboratory test values, and the proportion of discontinued subjects during the double-blind period. AEs were graded for severity according to the Common Terminology Criteria for Adverse Events (CTCAE) version 5·0 and relation to the study drug. The secondary outcomes for the feasibility trial objective included the ratio of the change in the ALSFRS-R score every 4 weeks between pre- and posttreatment assessments, change in the ALSFRS-R score,^12^ combined assessment of function and survival (CAFS) scores,^13^ amount of physical activity,^14,18^ composite endpoint as a sum of the Z-transformed scores of items (full items are provided in the protocol),^19^ time to death or a specified state of disease progression, and time to %FVC of ≤50%. Exploratory outcomes are provided in the protocol.

### 2.6 Statistical analysis

The probability of detecting an AE with a true incidence rate of 10% was 80% with 15 subjects in the treatment group. Additionally, five subjects were randomly assigned to the placebo group to collect efficacy and safety data and eliminate bias.

Statistical analyses and reporting of this trial were conducted in accordance with the CONSORT statement guidelines, with the primary analyses based on the intention-to-treat principle. For primary safety analysis, the proportion of AEs was estimated in each treatment group, and the exact 95% confidence interval (CI) was calculated by the binomial distribution.

The secondary outcomes for efficacy were analysed using an analysis-of-covariance (ANCOVA) model with fixed-effect terms for the study group and the corresponding baseline value as a covariate. The change-from-baseline means are the least-squares means from the ANCOVA model during the 24-week treatment period. For the sensitivity analysis, mean changes from baseline were analysed using a restricted maximum likelihood (REML)-based repeated measures approach in combination with the Newton Raphson algorithm. The mixed model for repeated measures (MMRM) analyses included the fixed, categorical effects of treatment, visit, and treatment by visit interaction. A common unstructured covariance structure was used for modelling within the patient error. If this analysis failed to converge, the following structures were tested until model convergence was achieved: Toeplitz, autoregressive, or compound-symmetry structures. The Kenward-Roger approximation was used to estimate the denominator degrees of freedom. The calculation of risk difference and 95% CI according to the Newcombe-Wilson method was added to the efficacy endpoints. No adjustments were made for the multiple testing of secondary outcomes because of the exploratory nature of the study. Missing values in outcome variables were not imputed because mixed models can handle missing data by maximum likelihood.

For time to event, the Kaplan–Meier method was applied to assess overall survival, and competing risk analyses using the Cox model for time to death or a specified state of disease progression and the Fine and Grey model for time to %FVC of ≤50% were performed to assess disease progression times. Cumulative incidence curves are presented for each treatment group. All comparisons were planned, and all p values are two-sided. A p value < 0·05 was considered statistically significant. All statistical analyses were performed using SAS software version 9·4 (SAS Institute Inc., Cary, NC, USA) and described in the statistical analysis plan, which was fixed prior to database locking. The trial was reported to the Japanese Pharmaceuticals and Medical Devices Agency (PMDA) (No. 2020-7717) and prospectively registered with the UMIN Clinical Trials Registry (number UMIN000034954) on November 30, 2018.

### Role of the funding source

The role of GSK was in providing ropinirole extended-release tablets and placebo and that of K Pharma was in funding.

## 3 RESULTS

### 3.1 characteristics of participants

From December 3, 2018, to September 12, 2019, we screened 29 participants, of whom 21 completed the run-in period and were randomized. Before drug administration, 1 participant withdrew due to protocol deviation. Among the remaining 20 participants, 13 received ropinirole, and 7 received placebo. Of the 18 participants who completed the last dose at week 24 in the double-blind period, 8 completed follow-up visits 24 days after the last dose at week 48 in the open-label active extension period (figure 1).

**Figure 1.**
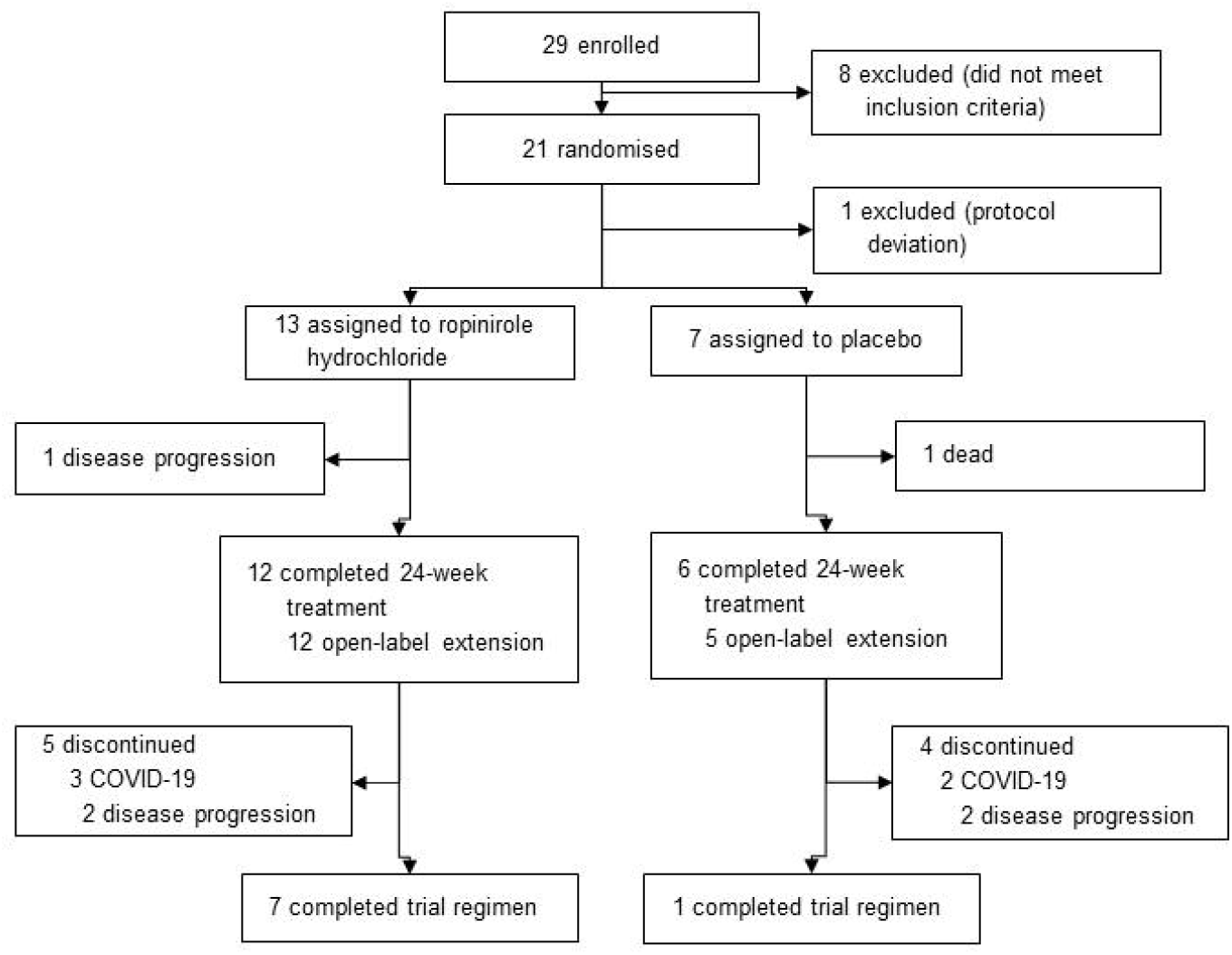
CONSORT Flow Diagram. The diagram illustrates the screening, randomization, and follow-up processes in the ROPALS trial. Trial participants who completed the run-in period were randomized to receive either ropinirole or placebo at a 3:1 ratio. Due to dynamic allocation and allocation adjustment factors, 13 participants were assigned to receive ropinirole and 7 participants were assigned to receive placebo (approximately a 2:1 ratio). The *in vitro* evaluations of drug effects and exploration of new biomarkers using patient iPSC-derived motor neurons are ongoing.

This was a small-scale phase 1/2a randomized control trial, and two more subjects were allocated to the placebo group than planned; therefore, the number of subjects in the placebo group was seven with an allocation ratio of 2:1. According to Kang et al, at the initial stages of a clinical study or for trials with small sample sizes, using simple randomization can cause an extreme imbalance among the treatment groups by random chance alone.^20^

Table 1 summarizes the demographic and baseline characteristics of the 20 participants who were randomized and received treatments. All participants were diagnosed with sporadic ALS with no detectable known pathogenic mutations in ALS causative genes (supplementary method). Both treatment groups had similar yet high proportions of participants with ≤grade 2 ALS severity. The ropinirole group had a longer disease duration (median: 24 months) than the control group (median: 17 months). Nevertheless, the total ALSFRS-R scores at pre-observation and baseline were balanced in both groups.

**Table 1.** Demographic and Clinical Characteristics of the 20 Participants at Baseline.*

|  | Ropinirole<br>hydrochloride | Placebo | Overall |
| --- | --- | --- | --- |
|  | (N=13) | (N=7) | (N=20) |
| Female sex — no. (%) | 3 (23·1) | 4 (57·1) | 7 (35·0) |
| Age — yr | 65·2±12·6 | 66·3±7·5 | 65·6±10·9 |
| Familial history — no. (%) | 0 (0) | 0 (0) | 0 (0) |
| Revised El Escorial Diagnostic criteria — no. (%) |  |  |  |
| Clinically Definite ALS | 1 (7·7) | 1 (14·3) | 2 (10·0) |
| Clinically Probable ALS | 10 (76·9) | 6 (85·7) | 16 (80·0) |
| Clinically Probable ALS Laboratory<br>Supported | 2 (15·4) | 0 (0) | 2 (10·0) |
| ALS severity scale§ — no. (%) |  |  |  |
| Grade 1 | 4 (30·8) | 3 (42·9) | 7 (35·0) |
| Grade 2 | 8 (61·5) | 3 (42·9) | 11 (55·0) |
| Grade 4 | 1 (7·7) | 1 (14·3) | 2 (10·0) |
| Onset lesion — no. (%) |  |  |  |
| Bulbar | 4 (30·8) | 4 (57·1) | 8 (40·0) |
| Upper limb | 8 (61·5) | 1 (14·3) | 9 (45·0) |
| Lower limb | 1 (7·7) | 2 (28·6) | 3 (15·0) |
| Riluzole or edaravone use — no. (%) |  |  |  |
| Riluzole | 10 (76·9) | 5 (71·4) | 15 (75·0) |
| Edaravone | 6 (46·2) | 2 (28·6) | 8 (40·0) |
| Median months since ALS symptom onset†<br>(Interquartile range) | 24·0 (18-27) | 17·0 (6-18) | 18·0 (16-26) |
| Body-mass index□ | 22·91±3·82 | 19·69±2·63 | 21·78±3·72 |
| ALSFRS Total Score: Before observation¶ | 43·4±2·8 | 42·0±2·9 | 42·9±2·9 |
| ALSFRS Total Score: At baseline¶ | 40·0±2·9 | 38·6±3·1 | 39·5±3·0 |
| %FVC: At baseline — % | 94·4±14·9 | 81·4±23·2 | 89·9±18·7 |

\*Plus-minus values are the means ± SD. All participants were Japanese.
§According to the Japan ALS severity classification (grades 1–5, grade 5 is most severe).
¶ALSFRS-R consists of four subdomains and 12 components, where each component is scored on a scale from 0 to 4. Higher scores indicate better function.
□The body-mass index is the weight in kilograms divided by the square of the height in meters.
†The difference in disease duration resulted from the dynamic allocation of trial participants using “≥30 months or <30 months after onset” as an adjustment factor because we initially did not expect a statistically significant
- 1 variation within the group of participants. However, the group of participants in the <30 months after onset group - 2 was diverse. Abbreviations: ALS = Amyotrophic Lateral Sclerosis, ALSFRS-R = Revised ALS Functional Rating Scale, SD = Standard Deviation.

### 3.2 primary outcome

For the safety analyses, AEs and anticipated AEs were reported for all 20 participants in the double-blind period and 17 participants in the open-label extension period (table 2A, B).

**Table 2A.** Summary of Adverse Events* for 20 Participants.

|  | Double-blind period |  | Open-label extension period |  |
| --- | --- | --- | --- | --- |
|  | Ropinirole<br>hydrochloride | Placebo | Ropinirole<br>hydrochloride | Placebo<br>(Ropinirole<br>hydrochloride) |
| Variable | (N=13) | (N=7) | (N=12) | (N=5) |
| **Adverse event** |  |  |  |  |
| ≥1 Adverse event — no. (% , 95%CI) | 12 (92.3, 64.0-99.8) | 6 (85.7, 42.0-99.6) | 11 (91.7, 61.5-99.8) | 4 (80.0, 28.4-99.5) |
| No. of distinct events | 50 | 14 | 34 | 32 |
| Trial regimen interrupted owing to adverse event — no. (%) | 0 | 0 | 0 | 0 |
| Dose reduced owing to adverse event — no. (%) | 0 | 0 | 0 | 0 |
| Trial regimen discontinued owing to adverse event — no. (%) | 0 | 0 | 0 | 0 |
| **Serious adverse events** |  |  |  |  |
| ≥1 Adverse event — no. (%) | 0 | 1 (14.3) | 0 | 1 (20.0) |
| No. of distinct events | 0 | 1 | 0 | 1 |
| Death — no. (%) | 0 | 1 (14.3) | 0 | 0 |
| ≥1 Serious adverse event considered to be related to intervention — no. (%) | 0 | 0 | 0 | 1 (20.0) |
| Trial regimen discontinued owing to serious adverse event — no. (%) | 0 | 0 | 0 | 0 |
| **Adverse events with ≥5% incidence in either group† — no. (% , 95%CI)** |  |  |  |  |
| Gastrointestinal disorders | 10 (76.9, 46.2-95.0) | 1 (14.3, 0.4-57.9) | 1 (8.3, 0.2-38.5) | 3 (60.0, 14.7-94.7) |
| Musculoskeletal and connective-tissue disorders | 1 (7.7, 0.2-36.0) | 1 (14.3, 0.4-57.9) | 0 (0.0 - 26.5) | 0 (0.0-52.2) |
| Injury, poisoning, and procedural complications | 1 (7.7, 0.2-36.0) | 2 (28.6, 3.7-71.0) | 3 (25.0, 5.5-57.2) | 0 (0.0-52.2) |
| Nervous-system disorders | 7 (53.8, 25.1-80.8) | 0 (0.0-41.0) | 7 (58.3, 27.7-84.8) | 2 (40.0, 5.3-85.3) |
| Infections and infestations | 3 (23.1, 5.0-53.8) | 2 (28.6, 3.7-71.0) | 5 (41.7, 15.2-72.3) | 3 (60.0, 14.7-94.7) |
| Respiratory, thoracic, and mediastinal disorders | 1 (7.7, 0.2-36.0) | 1 (14.3, 0.4-57.9) | 0 (0.0 - 26.5) | 0 (0.0-52.2) |
| General disorders and administration-site conditions | 4 (30.8, 9.1-61.4) | 0 (0.0-41.0) | 4 (33.3, 9.9-65.1) | 3 (60.0, 14.7-94.7) |
| Skin and subcutaneous-tissue disorders | 4 (30.8, 9.1-61.4) | 0 (0.0-41.0) | 2 (16.7, 2.1-48.4) | 3 (60.0, 14.7-94.7) |
| Psychiatric disorders | 1 (7.7, 0.2-36.0) | 0 (0.0-41.0) | 0 (0.0 - 26.5) | 1 (20.0, 0.5-71.6) |
| Hepatobiliary disorders | 0 (0.0-24.7) | 0 (0.0-41.0) | 0 (0.0 - 26.5) | 1 (20.0, 0.5-71.6) |
| Renal and urinary disorders | 0 (0.0-24.7) | 0 (0.0-41.0) | 0 (0.0 - 26.5) | 1 (20.0, 0.5-71.6) |
| Metabolism and nutrition disorders | 0 (0.0-24.7) | 1 (14.3, 0.4-57.9) | 1 (8.3, 0.2-38.5) | 1 (20.0, 0.5-71.6) |
| Cardiac disorders | 0 (0.0-24.7) | 1 (14.3, 0.4-57.9) | 0 (0.0 - 26.5) | 0 (0.0-52.2) |
| Vascular disorders | 1 (7.7, 0.2-36.0) | 1 (14.3, 0.4-57.9) | 1 (8.3, 0.2-38.5) | 1 (20.0, 0.5-71.6) |
| Blood and lymphatic system disorders | 0 (0.0-24.7) | 0 (0.0-41.0) | 0 (0.0 - 26.5) | 1 (20.0, 0.5-71.6) |
| Eye disorders | 1 (7.7, 0.2-36.0) | 0 (0.0-41.0) | 1 (8.3, 0.2-38.5) | 0 (0.0-52.2) |
| Reproductive system and breast disorders | 0 (0.0-24.7) | 0 (0.0-41.0) | 1 (8.3, 0.2-38.5) | 0 (0.0-52.2) |
| Investigations | 0 (0.0-24.7) | 1 (14.3, 0.4-57.9) | 3 (25.0, 5.5-57.2) | 1 (20.0, 0.5-71.6) |

\*The safety population included all participants who received at least one dose of ropinirole or placebo. The relevance of adverse events or serious adverse events to the intervention was determined by the site investigator. †Adverse events and serious adverse events were classified according to system organ class and preferred terms in the *Medical Dictionary for Regulatory Activities*, version 23.1.

**Table 2B.** Summary of Anticipated Adverse Events* for 20 Participants.

|  | Double-blind period |  | Open-label extension period |  |
| --- | --- | --- | --- | --- |
| Variable | Ropinirole hydrochloride<br>(N=13) | Placebo<br>(N=7) | Ropinirole hydrochloride<br>(N=12) | Placebo (Ropinirole hydrochloride)<br>(N=5) |
| **Anticipated adverse event** |  |  |  |  |
| ≥1 Adverse event — no. (% , 95%CI) | 9 (69.2, 38.6-90.9) | 1 (14.3, 0.4-57.9) | 8 (66.7, 34.9-90.1) | 2 (40.0, 5.3-85.3) |
| No. of distinct events | 18 | 1 | 11 | 9 |
| Trial regimen interrupted owing to adverse event — no. (%) | 0 | 0 | 0 | 0 |
| Dose reduced owing to adverse event — no. (%) | 0 | 0 | 0 | 0 |
| Trial regimen discontinued owing to adverse event — no. (%) | 0 | 0 | 0 | 0 |
| **Anticipated serious adverse events** |  |  |  |  |
| ≥1 Adverse event — no. (%) | 0 | 0 | 0 | 1 (20.0) |
| No. of distinct events | 0 | 0 | 0 | 1 |
| Death — no. (%) | 0 | 0 | 0 | 0 |
| ≥1 Serious adverse event considered to be related to intervention — no. (%) | 0 | 0 | 0 | 1 (20.0) |
| Trial regimen discontinued owing to serious adverse event — no. (%) | 0 | 0 | 0 | 0 |
| **Anticipated adverse events with ≥5% incidence in either group† — no. (% , 95%CI)** |  |  |  |  |
| Gastrointestinal disorders | 6 (46.2, 19.2-74.9) | 0 (0.0-41.0) | 1 (8.3, 0.2-38.5) | 1 (20.0, 0.5-71.6) |
| Musculoskeletal and connective-tissue disorders | 0 (0.0-24.7) | 0 (0.0-41.0) | 0 (0.0-26.5) | 0 (0.0-52.2) |
| Injury, poisoning, and procedural complications | 0 (0.0-24.7) | 0 (0.0-41.0) | 0 (0.0-26.5) | 0 (0.0-52.2) |
| Nervous-system disorders | 3 (23.1, 5.0-53.8) | 0 (0.0-41.0) | 3 (25.0, 5.5-57.2) | 2 (40.0, 5.3-85.3) |
| Infections and infestations | 0 (0.0-24.7) | 0 (0.0-41.0) | 0 (0.0-26.5) | 0 (0.0-52.2) |
| Respiratory, thoracic, and mediastinal disorders | 1 (7.7, 0.2-36.0) | 0 (0.0-41.0) | 0 (0.0-26.5) | 0 (0.0-52.2) |
| General disorders and administration-site conditions | 3 (23.1, 5.0-53.8) | 0 (0.0-41.0) | 3 (25.0, 5.5-57.2) | 2 (40.0, 5.3-85.3) |
| Skin and subcutaneous-tissue disorders | 0 (0.0-24.7) | 0 (0.0-41.0) | 1 (8.3, 0.2-38.5) | 0 (0.0-52.2) |
| Psychiatric disorders | 0 (0.0-24.7) | 0 (0.0-41.0) | 0 (0.0-26.5) | 0 (0.0-52.2) |
| Hepatobiliary disorders | 0 (0.0-24.7) | 0 (0.0-41.0) | 0 (0.0-26.5) | 0 (0.0-52.2) |
| Renal and urinary disorders | 0 (0.0-24.7) | 0 (0.0-41.0) | 0 (0.0-26.5) | 0 (0.0-52.2) |
| Metabolism and nutrition disorders | 0 (0.0-24.7) | 0 (0.0-41.0) | 0 (0.0-26.5) | 0 (0.0-52.2) |
| Cardiac disorders | 0 (0.0-24.7) | 0 (0.0-41.0) | 0 (0.0-26.5) | 0 (0.0-52.2) |
| Vascular disorders | 0 (0.0-24.7) | 0 (0.0-41.0) | 0 (0.0-26.5) | 0 (0.0-52.2) |
| Blood and lymphatic system disorders | 0 (0.0-24.7) | 0 (0.0-41.0) | 0 (0.0-26.5) | 1 (20.0, 0.5-71.6) |
| Eye disorders | 0 (0.0-24.7) | 0 (0.0-41.0) | 0 (0.0-26.5) | 0 (0.0-52.2) |
| Reproductive system and breast disorders | 0 (0.0-24.7) | 0 (0.0-41.0) | 1 (8.3, 0.2-38.5) | 0 (0.0-52.2) |
| Investigations | 0 (0.0-24.7) | 1 (14.3, 0.4-57.9) | 1 (8.3, 0.2-38.5) | 0 (0.0-52.2) |

\*The safety population included all participants who received at least one dose of ropinirole or placebo. The relevance of adverse events or serious adverse events to the intervention was determined by the site investigator.
†Adverse events and serious adverse events were classified according to system organ class and preferred terms in the *Medical Dictionary for Regulatory Activities*, version 23.1.

During the double-blind period, one death occurred in the placebo group (idiopathic ischaemic heart disease on day 157), which was attributed to disease progression by the investigators. No participant discontinued treatment due to adverse experiences in either treatment group. In total, 92·3% (12/13) and 85·7% (6/7) of participants experienced at least one AE in the ropinirole group and the placebo group, respectively. AEs that occurred in ≥10% of participants in both treatment groups included gastrointestinal disorders (14·3% for placebo and 76·9% for ropinirole) and infection and infestations (28·6% for placebo and 23·1% for ropinirole). Ropinirole-related AEs occurring in ≥20% of participants included constipation (61·5%), nausea (38·5%), somnolence (30·8%), and headache (23·1%). No statistically significant abnormalities were detected for conventional clinical laboratory measurements or CSF tests.

During the open-label active extension period, at least one AE occurred in 91·7% of participants in the ropinirole-ropinirole (RR) treatment group and 80% of participants in the placebo-ropinirole (PR) treatment group. The highest incidence of AEs (≥20% in any treatment group) was related to nervous-system disorders (40% in the PR group and 58·3% in the RR group), infections and infestations (60% in the PR group and 41·7% in the RR group), and general disorders and administration-site conditions (60% in the PR group and 33·3% in the RR group). Extended ropinirole treatment was associated with a higher frequency of somnolence (40%) and constipation (60%). Pharmacokinetic data are summarized in figure S2 and table S1.

### 3.3 secondary outcomes

#### 3.3.1 Functional outcomes

During the double-blind period, the mean ALSFRS-R slope per month before and after treatment was 1·042 (95% CI, 0·249 to 1·835) in the ropinirole group compared with 1·254 (95% CI, 0·165 to 2·344) in the placebo group (table S2). The difference between groups was −0·212 (95% CI, −1·577 to 1·153). The change in the ALSFRS-R score from day 1 to week 24 of the double-blind period was −5·36 (95% CI, −8·09 to −2·64) for the ropinirole group and −6·82 (95% CI, −10·54 to −3·10) for the placebo group (table S3). The between-group difference in the change in ALSFRS-R scores was 1·46 points (95% CI, −3·15 to 6·07) over 24 weeks of treatment (figure 2A). In addition, participants in the ropinirole group had higher daily physical activity, as measured by METs (figure 2B). The difference in METs per month between treatment groups at week 24 was 317·5 (95% CI, 66·3 to 568·7) (table S3). There was a persistent increase in between-group differences in the change in ALSFRS-R scores beyond 24 weeks. The change in the ALSFRS-R score during the entire treatment period was −7·64 (95% CI, −10·66 to −4·63) for the RR group and −17·51 (95% CI, −22·46 to −12·56) for the PR group (table S3). The treatment difference in the change in ALSFRS-R scores was 9·86 points (95% CI, 4·07 to 15·66) over 48 weeks, indicating that RR reduced functional decline (figure 2A). The ALSFRS-R score for each of the 12 items is also shown (figure S3). Furthermore, we applied a composite endpoint that combined multiple clinically relevant functional components, expressed as the mean z-score. The between-group differences in the mean z-score were 11·27 (95% CI, −5·85 to 28·39), 25·04 (95% CI, 4·34 to 45·74), and 15·27 (95% CI, −11·92 to 42·47) at the end of 24, 39, and 50 weeks, respectively (figure 2C). The results of individual components within the composite endpoint are shown in Figures S4–S7 and Tables S4–S7.

**Figure 2.**
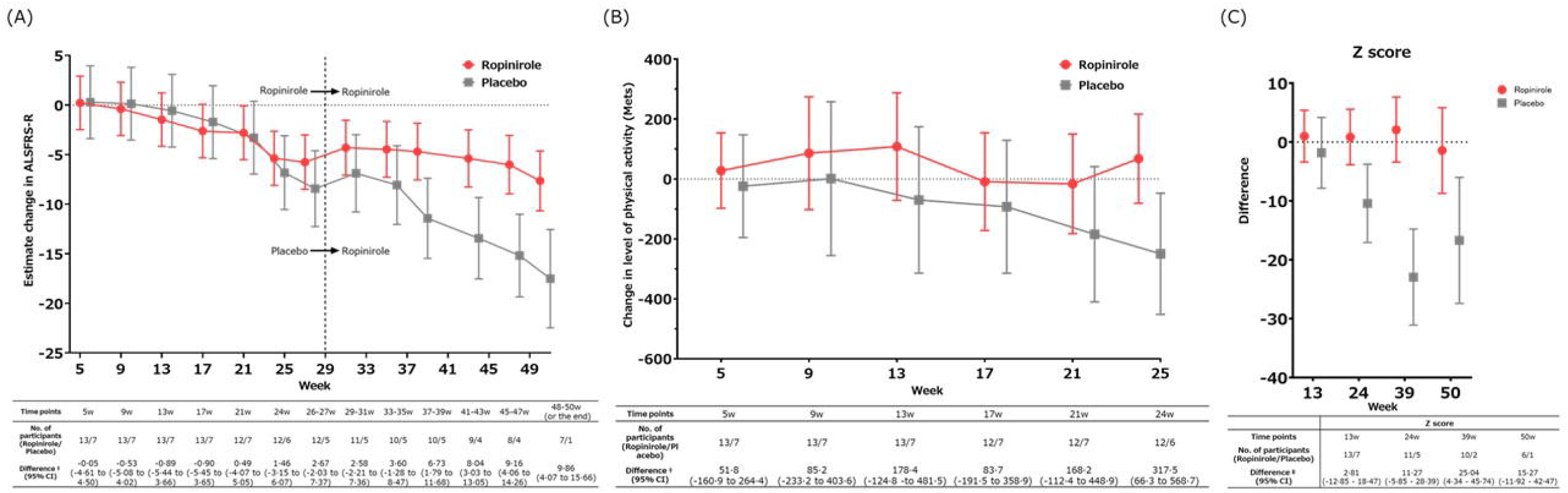
Effect of Ropinirole Treatment on ALSFRS-R score, Physical Activity and Z-score. (A) Estimated change from baseline in ALSFRS-R total score every four weeks (FAS, entire period). (B) Level of physical activity every four weeks (FAS, double-blind period). (C) The composite z-score functional outcome based on a battery of functional and QoL endpoints (FAS, entire trial period; post hoc). → Z: The amount of change at each measurement time (weeks 13, 24, 39, 50) from the values at 12 weeks after interim registration in each item was calculated, and then converted to a z-score. Next, a comparison between groups was performed. ‡Differences >0 indicate a positive treatment effect of ropinirole. I bars represent the 95%CI. Abbreviations: ALSFRS-R = Revised ALS Functional Rating Scale, FAS = Full Analysis Set,

#### 3.3.2 Survival outcomes

To examine the relevance of the ALSFRS-R benefit to survival, we used the CAFS measurement, which adjusts the ALSFRS-R score against mortality. The CAFS score favoured ropinirole only in the open-label extension period (difference in median CAFS scores, 6·0; 95% CI, −5 to 9) and the entire treatment period (difference in median CAFS scores, 9·0; 95% CI, 1 to 12) but not in the double-blind period (difference of median CAFS scores, 4·0; 95% CI, −6 to 10) (figure 3A). Related to respiratory outcome, 14·3% of participants in the placebo group reached ≤50%FVC in contrast to 0% in the ropinirole group in the double-blind period (95% CI, −51·3 to 11·4), while 50% of participants in the PR group reached ≤50%FVC beyond 40 weeks in contrast to 0% in the RR group (RR group: 7·7%; PR group: 42·9%; difference, −35·2; 95% CI, −67·9 to 2·1) (figure 3B). Notably, %FVC at baseline was not different between the RR and RP groups (95% CI, −4·85 to 30·8).

**Figure 3.**
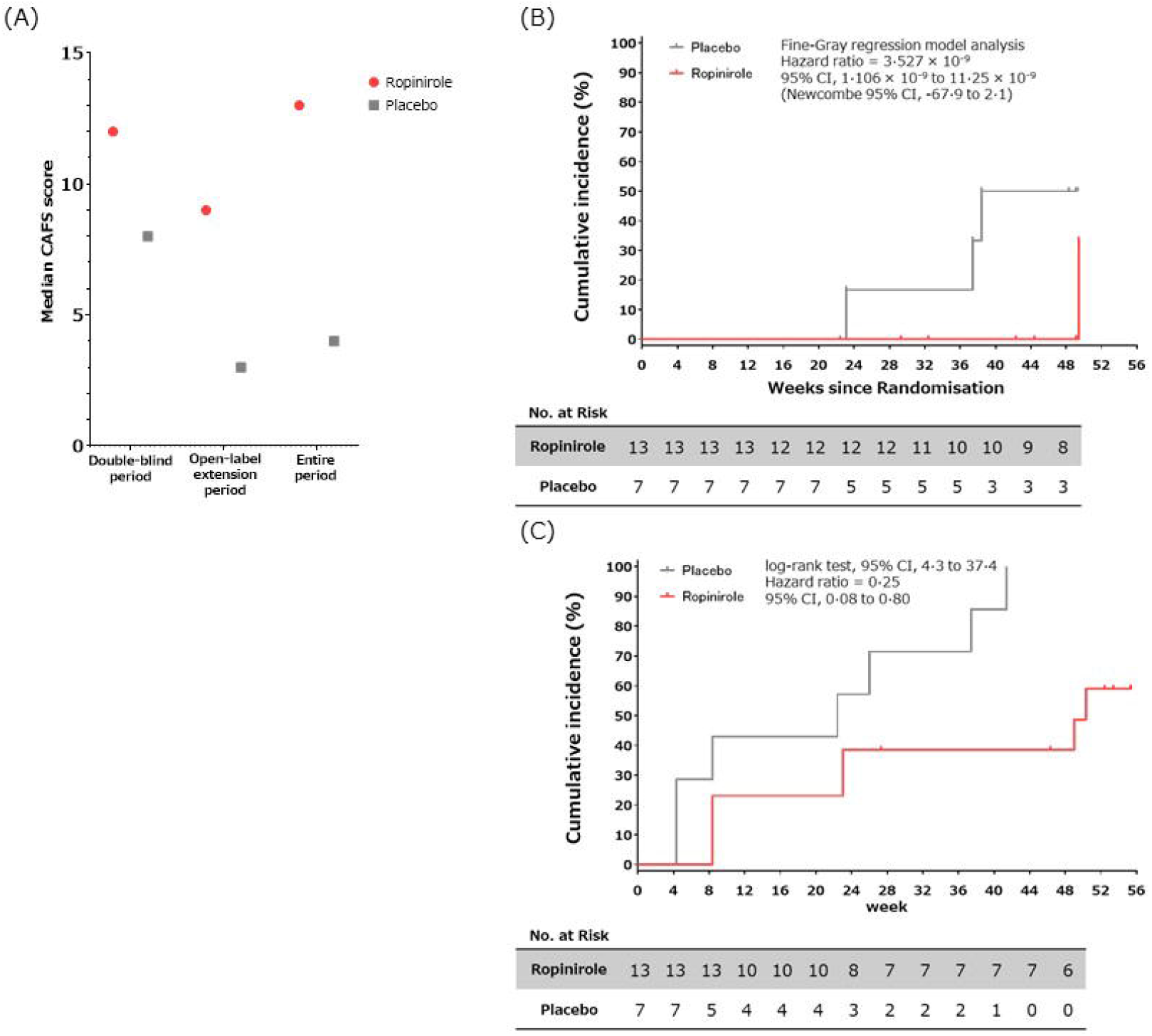
Effect of Ropinirole Treatment on Survival. (A) Median CAFS scores^¶^ of participants in the double-blind period (n=20), open-label period (n=16), and entire trial period (n=20). Scores range from (1) (worst) to (20) (best). (B) The proportion of participants whose %FVC was reduced to 50% over 48 weeks. (C) The proportion of participants with an occurrence of disease progression events, including death, over 48 weeks. ^¶^CAFS ranks patient clinical outcomes based on survival time and change in the ALSFRS-R score. Each patient’s outcome is compared with every other patient’s outcome, assigned a score, and the summed scores are ranked. The mean rank score for each treatment group is then calculated. A higher mean CAFS score indicates a better group outcome. Abbreviations: CAFS = Combined Assessment of Function and Survival, %FVC = percent predicted Forced Vital Capacity, ALSFRS-R = Revised ALS Functional Rating Scale.

We also investigated the time to death or certain disease progression events. There were 7 out of 7 (100%) events in the PR group and 7 out of 13 (54%) events in the RR group, suggesting a twofold decrease in disease progression in the RR group (tables S9, S10A). The RR group had an extended time to the first disease progression event (median event-free survival: 50·3 weeks vs. 22·4 weeks in the PR group) (log-rank test: 95% CI, 4·3 to 37·4; Cox regression model analysis: hazard ratio = 0·25, 95% CI, 0·08 to 0·80) (figure 3C). We observed a higher proportion of participants who progressed to grade 4 ALS severity or above in the placebo group at the end of 24 weeks (71% for placebo and 53% for ropinirole groups) and 48 weeks (100% for PR and 58% for RR groups) (figure S8, table S9B). Moreover, we identified liquid biomarkers related to the pathomechanism of ALS (figure S9A–C).

## 4 DISCUSSION

In our previous *in vitro* drug efficacy study using patient-derived iPSC-MNs^5^, the effect of ropinirole on ALS was evident at 0·1, 1, and 10 µmol/L. The estimated ropinirole concentration at 2 mg of Requip CR is approximately 5 nmol/L and that at 16 mg is approximately 50 nmol/L in plasma and CSF.^21^ In other experiments, the efficacy of ropinirole was dose-dependent at ropinirole concentrations of 1 nmol/L and 10 µmol/L (figure S10), indicating that 2 mg of Requip CR is effective. However, it is desirable to administer up to 16 mg, which is the maximum approved dosage for use in PD in Japan.

Following the administration of 16 mg of ropinirole, the CSF concentration of ropinirole reached approximately 18 nmol/L (table S1) and remained at a steady state. This observation is consistent with the effective concentration of ropinirole in suppressing ALS-related damage in iPSC-derived MNs, which ranged from 1 nmol/L or more based on the present (figure S10) and previous results.^5^ Steady exposure to 18 nmol/L ropinirole was not associated with any serious AEs that led to trial discontinuation. This trial demonstrated that fixed-dose ropinirole (16 mg) had no specific safety concerns.

Overall, the incidence of AEs, most of which had been reported previously, were similar within both groups (table 2A, B). Although gastrointestinal disorders had a high incidence at 76·9% in the ropinirole group (14·3% in the placebo group), these symptoms were temporary mild nausea and diarrhoea, and there were no participants in which the continuation of oral administration was hindered (table 1). Therefore, ropinirole (2–16 mg) is tolerable for ALS patients.

Interestingly, the incidence of ropinirole-induced gastrointestinal disorders in PD patients and healthy individuals in Japan is approximately 5-12% (pharmaceutical interview form for Requip CR tablets, version 6) and 0% (table S10), respectively, which is different from the results in ALS patients in this study. Therefore, gastrointestinal symptoms are not considered to be a major adverse effect of ropinirole, and the reason why many gastrointestinal symptoms appeared in ALS patients in this clinical trial is unknown. Regarding safety concerns, gastrointestinal adverse effects for ALS patients might complicate care because ALS patients have poor BMI, malnutrition, and cachexia; however, the ropinirole group did not differ significantly from the placebo group in weight drop or in the ALSAQ-40 (eating and drinking) score (figures S6 and S7). Thus, we thought that gastrointestinal symptoms did not significantly affect safety or tolerability in this trial. Because the study was primarily designed to assess safety and tolerability, we chose to present our clinical findings regarding the secondary outcomes as reference for further study. Moreover, regarding the assurance of blinding, the ALSFRS-R score includes both subjective and objective assessments. The investigators did indeed evaluate the patients objectively, assessing at least half of the ALSFRS-R items (i.e., speech, handwriting, cutting food and handling utensils, walking, climbing stairs and respiratory insufficiency). In our posthoc analysis of the transition of each of the 12 items in the ALSFRS-R, no obvious differences were noted between the results of subjective and objective assessment items (figure S3). Therefore, blinding was not broken, and outcomes were not influenced throughout the study. The blinding was kept throughout, and it was plausible that patients and investigators would not know which group had active or placebo treatment; therefore, outcomes were not influenced during the study.

The estimated effect size of ropinirole observed for a change in the ALSFRS-R score ranged from 1·46 points to 9·86 points over 48 weeks, equivalent to a 21%–60% slower rate of functional decline. Neurologists generally perceive a treatment difference of >20% as somewhat clinically meaningful.^22^ Accordingly, the FDA and PMDA-approved ALS drug edaravone reduced the rate of functional decline by 33% over 24 weeks.^23^ Moreover, clinical evaluation items such as the CAFS score, muscle strength, physical activity, pulmonary function, and composite z-score might be useful as evaluation indexes for the efficacy of ropinirole. The CAFS endpoint is advantageous because it evaluates functional and survival outcomes concomitantly without relying on statistical assumptions. CAFS is well adjusted for missing values due to death or drop-outs.^13^ However, CAFS measurements may be less relevant because there was only one death in this study, indicating that mortality did not confound the survival analysis *per se*. Ropinirole, a non-ergot dopamine agonist, was previously approved for the treatment of PD. The full mechanism of action of ropinirole in ALS is not yet understood. Based on our previous reports and according to its structural characteristics, its effects might be dopamine D2 receptor (D2R)-dependent and D2R-independent.^5, 21^ With regard to the dopamine D2R-dependent anti-ALS mechanism of ropinirole, we confirmed the protein expression of dopamine D2R on anterior horn cells in a healthy human spinal cord and human iPSC-derived LMNs (figures S11 and S12), which is consistent with the transcriptome data.^21^ Furthermore, D2R couples with Gi proteins to inhibit adenyryl cyclase, resulting in decreased intracellular cAMP levels. As a result, neuronal hyperexcitability, which is thought to be the cause of MN toxicity, can be inhibited.^24,25^ Recently, it was shown that D2R agonists other than ropinirole, such as bromocriptine and sumanirole, suppressed the hyperexcitability of human iPSC-derived MNs *in vitro*.^24^ This suppression of neuronal hyperexcitation might protect ALS MNs, which is relevant to the action of riluzole. Furthermore, previous reports indicated that autophagy may be activated by dopamine D2R and D3R agonists via a Beclin-1-dependent pathway.^26, 27^ Thus, ropinirole-induced dopamine D2R activation might induce autophagy, supporting the degradation and disassembly of abnormal RNA-protein complexes in the MNs of patients with ALS.

With regard to dopamine D2R-independent mechanisms, it is notable that ropinirole is a lipophilic cation that readily localizes to the mitochondrial inner membrane due to its tertiary amine moiety and possesses antioxidant properties related to its oxindole structure.^21^ Consistent with this, urine 8-hydroxy-2’-deoxyguanosine, a surrogate marker of oxidative stress, was not increased in the ropinirole group throughout the entire study period (figure S9A). These findings suggest that ropinirole may scavenge mitochondrial reactive oxygen species and protect cells against mitochondrial damage in ALS. In addition, ropinirole suppressed increases in ALS pathomechanism-related markers, such as serum ferritin^28, 29^ and high-sensitivity C-reactive protein,^30^ throughout the entire study period (figure S9A). Interestingly, bromocriptine mesylate, an ergot alkaloid and agonist of the same dopamine receptor, namely, D2R, is expected to have an effect on dopamine D2R-independent and neuronal apoptosis inhibitory protein (NAIP), an oxidative stress-induced cell death suppressor -mediated anti-ALS mechanism.^6,31^ Collectively, the precise mechanism of action of ropinirole in ALS warrants further investigation.

Interpretations of efficacy analyses in this feasibility study were limited by the small sample size of 20 participants. This was further compounded by the unexpectedly higher rate of discontinuation in this study than the historical rate in clinical trials of ALS (∼20%)^32^;46·2% (6/13) of the participants in the ropinirole group and 85·7% (6/7) in the placebo group discontinued the trial (table S11), and the ratio of patients in the ropinirole versus placebo group changed, particularly in the open-label extension period.

These discontinuation rates, particularly during the open-label extension phase, were attributable, at least in part, to the COVID-19 outbreak, which accounted for 23% and 29% of the participants in the ropinirole and placebo groups, respectively. Therefore, it may have had some undesirable influence on the objective interpretation of the results. The number of participants in the placebo group who discontinued the trial due to a worsening condition (47%) was higher than that in the ropinirole group (23%). We did not apply any imputation to the missing data but used the MMRM to infer missing data. In a recent randomized and controlled trial of rasagiline for ALS, 60% of the placebo group withdrew from the trial. As a result, the investigators increased the power of the study by utilizing placebo (n=8) and historical placebo controls (n=177).^33^ In the future, the addition or adoption of historical placebo controls may be a plausible approach to enrich the placebo cohort, particularly for trials with an extended period.

In this trial, the placebo group had more bulbar onset, more female patients and a lower BMI at baseline, which could be the relevant prognostic factors for ALS.^34-36^ These are major weaknesses of this trial. In addition, there are some points to be discussed for further study. It is important to note that the slope for the ALSFRS-R decline (−3·4 points) was consistent between the ropinirole and placebo groups during the 12-week run-in period. Additionally, the onset of disease is self-reported and can be influenced by the sensitivity and character of an individual patient; therefore, it is difficult to confirm the accuracy of the period of disease duration. We considered the latest disease progression rate to be more important and emphasized this progression rate with the ALSFRS-R during the 12-week pre-observation period. These observations ruled out potential baseline imbalances related to differences in disease duration. Moreover, previous observational studies showed that the relationship between bulbar onset and survival prognosis were controversial after adjustments for the other clinical features of ALS. ^34-37^ In practice, both the bulbar-onset group and the nonbulbar-onset group showed almost identical transitions in ALSFRS-R scores. In addition, both the ropinirole and placebo groups were divided into the bulbar-onset group and the nonbulbar-onset group (figure S13). When we compared the bulbar-onset and the nonbulbar-onset groups, no significant difference was observed at any time point. Therefore, in this clinical trial, the transition in the ALSFRS-R was not affected by the higher percentage of bulbar-onset patients in the placebo group.

Although we observed clinically meaningful differences between the treatment groups for functional and survival outcomes, these differences were primarily observed in the open-label phase. During the double-blind phase, we observed statistically significant treatment differences only for physical activity. Indeed, the rate of decline in the ALSFRS-R score in the placebo group was not improved even during the open-label extension period when the active drug was started. Interestingly, a similar phenomenon was observed in another trial.^38^ Although the underlying mechanism of this observation remains unclear, we postulate that taking ropinirole earlier and for longer is necessary to demonstrate its efficacy in participants with ALS. Furthermore, the patient population was skewed towards early-stage ALS (the total ALSFRS-R score at baseline was 39·5±3·0); thus, we also have no conclusive empirical data to demonstrate whether ropinirole was effective in patients with advanced ALS. Taken together, the uncontrolled data with a limited sample size reported in this feasibility study should be interpreted with caution and must be further validated with, ideally, advanced trials and trial designs. Therefore, the efficacy of ropinirole in ALS deserves further investigation in larger, multinational, randomized controlled trials.^39^

Although this trial is a randomized feasibility trial, the results of this trial encourage further study using ropinirole in ALS worldwide. To the best of our knowledge, the ROPALS trial is the representative touchstone of iPSC-based drug repurposing-enabled trials to define the feasibility of iPSC models in predicting clinical outcomes and replacing failure-prone preclinical transgenic mouse models of ALS.

## Supporting information

SECTION S1-3

SECTION S4-5

## Data Availability

All data produced in the present study are available upon reasonable request to the authors.

## 6 APPENDIX

### Funding

This clinical trial was sponsored by K Pharma, Inc. The study drug, active drugs, and placebo were supplied free-of-charge by GlaxoSmithKline K.K.

### Declaration of interests

Dr. Morimoto reports grants from Keio University School of Medicine, during the conduct of the study. Dr. Takahashi reports grants from Keio University School of Medicine, during the conduct of the study. Dr. Ito has nothing to disclose. Dr. Daté has nothing to disclose. Dr. Okada has nothing to disclose. Dr. Chai has nothing to disclose. Dr. Nishiyama has nothing to disclose. Dr. Naoki Suzuki has nothing to disclose. Dr. Fujimori reports personal fees from Sumitomo Dainippon Pharma Co., Ltd., outside the submitted work. Dr. Takao has nothing to disclose. Ms. Hirai has nothing to disclose. Dr. Kabe has nothing to disclose. Dr. Suematsu has nothing to disclose. Dr. Jinzaki has nothing to disclose. Dr. Aoki reports grants from Research on Nervous and Mental disorders, Research on rare and intractable diseases, Research on Psychiatric and Neurological Diseases and Mental Health from the Japanese Ministry of Health Labour and Welfare, Grants-in-Aids for Scientific Research, an Intramural Research Grant for Neurological Psychiatric Disorders from NCNP, Grants-in-Aids for Scientific Research from the Japanese Ministry of Education, Culture, Sports, Science and Technology (MEXT), and Practical Research Project for Rare / Intractable Diseases from Japan Agency for Medical Research and Development (AMED), during the conduct of the study. Mr. Fujiki has nothing to disclose. Dr. Sato has nothing to disclose. Dr. Suzuki has nothing to disclose. Dr. Nakahara reports grants from Japan Agency for Medical Research and Development (AMED), grants from K Pharma, Inc, non-financial support from GSK, during the conduct of the study. Dr. Okano reports grants from Japan Agency for Medical Research and Development, grants and personal fees from K Pharma Inc., during the conduct of the study; personal fees from SanBio Co.Ltd., personal fees from Regenerative Medicine iPS Gateway Centre Co.,Ltd., outside the submitted work; In addition, Dr. Okano has a patent THERAPEUTIC AGENT FOR AMYOTROPHIC LATERAL SCLEROSIS AND COMPOSITION FOR TREATMENT licensed to K Pharma Inc. and grant support from AMED (The Acceleration Program for Intractable Disease Research Utilizing Disease-specific iPS Cells (Grant No. JP 19bm0804003, JP 20bm0804003, JP 21bm0804003) and Research on Practical Application of Innovative Pharmaceutical and Medical Devices for Rare and Intractable Diseases (Grant No. JP 18ek0109395, JP 19ek0109395, JP 20ek0109395, JP 18ek0109329, JP 19ek0109329, JP 20ek0109329). The human spinal cord samples were provided by Platform of Supporting Cohort Study and Biospecimen Analysis, Grant-in-Aid for Scientific Research on Innovative Areas—Platforms for Advanced Technologies and Research Resources, Ministry of Education, Culture, Sports, Science and Technology, Japan (Grant No. 16H06277) and Intramural Research Grant for Neurological Psychiatric Disorders from National Centre of Neurology and Psychiatry (NCNP). The funding sources had no role in the analysis.

## Acknowledgments

We thank the trial coordinators, staff, and participants for their contributions, Komei Fukushima of K Pharma Inc, for contributing to the development of an earlier version of the research plan, Asuka Ogasawara, for coordinating this trial as one of the clinical research coordinators, Kazuo Watanabe of CTD Inc., a contract research organization, for managing this trial, Kentaro Higashi and Takashi Kasama of Keio University Hospital Clinical And Translational Research Center, for managing this trial as project managers, Takayuki Abe of Keio University Hospital Clinical And Translational Research Center, for contributing to the development of an earlier version of the statistical plan, Akihisa Yamazaki in the Department of Radiology, Keio University Hospital, for contributing to determining the muscle CT imaging conditions, Yoshitaka Yamada of the Department of Radiology, Keio University Hospital, for supporting the analysis of muscle CT images, Hiroyoshi Shiina in the Department of Pharmacy, Keio University Hospital, for contributing to managing the investigational drugs, Shun Kawada in the Office of Research Development and Sponsored Projects, Keio University School of Medicine for managing this trial as a contractor, Takanori Yokota of the Department of Neurology, Tokyo Medical and Dental University, Kazutomi Kanemaru, the Department of Neurology, Tokyo Metropolitan Geriatric Hospital, Masaki Takao, the Department of Clinical Laboratory, National Center of Neurology and Psychiatry (NCNP) for Independent Data Monitoring Committee, Fumiko Ozawa and Shiho Nakamura, for contributing to the *in vitro* analysis, and Mitsutoshi Tano at the Mihara Memorial Hospital, Isesaki, Japan for preparing and immunostaining the human spinal cord samples. We are grateful to Hitoshi Warita, Naoko Shimakura, Shion Osana, Ryo Funayama, Keiko Nakayama, Tetsuya Niihori, and Yoko Aoki, the Department of Neurology, Tohoku University Graduate School of Medicine, for their technical assistance on gene analysis, and BT Slingsby, Catalys Pacific, LLC for critical reading. We thank J. Ludovic Croxford, PhD, and Melissa Crawford, PhD, from Edanz (https://jp.edanz.com/ac), and Springer Nature for editing a draft of this manuscript.

## Data Sharing

The authors confirm that the data supporting the findings of this study are available within the article and its supplementary materials.

## Notes

### Clinical Trial

UMIN000034954

### Author Declarations

Ethics committee/IRB of Keio University Hospital gave ethical approval for this work (No. D18-01).

