## Supplementary material for "Ropinirole hydrochloride for amyotrophic lateral sclerosis: A single-center, randomized feasibility, double-blind, placebo-controlled trial": SECTION S1-3

### SUPPLEMENTARY APPENDIX

#### SECTION S1: STUDY INVESTIGATORS

| Name | Institution |
|---|---|
| Jin Nakahara | Department of Neurology, Keio University Hospital, Tokyo, Japan |
| Shinichi Takahashi | Department of Neurology, Keio University Hospital, Tokyo, Japan |
| Mamoru Shibata | Department of Neurology, Keio University Hospital, Tokyo, Japan |
| Shigeaki Suzuki | Department of Neurology, Keio University Hospital, Tokyo, Japan |
| Takahito Yoshizaki | Department of Neurology, Keio University Hospital, Tokyo, Japan |
| Daisuke Ito | Department of Neurology, Keio University Hospital, Tokyo, Japan |
| Yoshikane Izawa | Department of Neurology, Keio University Hospital, Tokyo, Japan |
| Morinobu Seki | Department of Neurology, Keio University Hospital, Tokyo, Japan |
| Kazutomi Kanemaru | Department of Neurology, Tokyo Metropolitan Geriatric Hospital and Institute of Gerontology, Tokyo, Japan |
| Naoko Makishi | Department of Neurology, Tokyo Metropolitan Geriatric Hospital and Institute of Gerontology, Tokyo, Japan |
| Rie Motoyama | Department of Neurology, Tokyo Metropolitan Geriatric Hospital and Institute of Gerontology, Tokyo, Japan |
| Satoru Morimoto | Department of Neurology, Tokyo Metropolitan Geriatric Hospital and Institute of Gerontology, Tokyo, Japan |
| Tomohiro Hikata | Department of Orthopaedic Surgery, Kitasato University Kitasato Institute Hospital, Tokyo, Japan |
| Wataru Takita | Department of Neurology, National Hospital Organization Nagoya Medical Center, Aichi, Japan |
| Keiji Kurita | Department of Neurology, Samukawa Hospital, Samukawa Shrine, Kanagawa, Japan |
| Ryoji Shimoyama | Department of Neurology, National Hospital Organization Matsue Medical Center, Shimane, Japan |
| Kazuhiro Itaya | Department of Neurology, Ichigao Hospital, Kanagawa, Japan |
| Hiroaki Yokote | Department of Neurology, Nitobe Memorial Nakano General Hospital, Tokyo, Japan |
| Kazuhiro Ishii | Department of Neurology, Faculty of Medicine, University of Tsukuba, Ibaraki, Japan |
| Asuka Yasuura | Department of Neurology, Japanese Red Cross Musashino Hospital, Tokyo, Japan |
| Takao Ienaga | Department of Neurology, Shin-Kuki General Hospital, Saitama, Japan |
| Ryo Usui | Department of Neurology, Kitasato University School of Medicine, Kanagawa, Japan |
| Ryotaro Ikeguchi | Department of Neurology, Tokyo Women's Medical University, Tokyo, Japan |
| Ryuichi Ohkubo | Department of Neurology, Fujimoto General Hospital, Miyagi, Japan |
| Makoto Nomura | Department of Neurology, Moriya Daiichi General Hospital, Ibaraki, Japan |
| Hiroaki Fujita | Department of Neurology, Dokkyo Medical University, Tochigi, Japan |
| Risa Kato | Department of Neurology, Kansai Medical University, Osaka, Japan |
| Yusuke Mochizuki | Department of Neurology, Hokushin General Hospital Nagano Prefectural Federation of |

| | |
|---|---|
| | Agricultural Cooperatives for Health and Welfare, Nagano, Japan |
| Masahiro Kato | Department of Neurology, Medical Plaza Edogawa, Tokyo, Japan |
| Ban Mihara | Department of Neurology, Institute of Brain and Blood Vessels, Mihara Memorial Hospital, Gunma, Japan |
| Hirokazu Yoshihashi | Department of Neurology, Kobari General Hospital, Chiba, Japan |
| Hironobu Myojin | Department of Neurology, Seirei Hamamatsu General Hospital, Shizuoka, Japan |
| Takeshi Yamada | Department of Neurology, Saiseikai Fukuoka General Hospital, Fukuoka, Japan |
| Yuichiro Furuya | Department of Neurology, National Defence Medical College Hospital, Saitama, Japan |
| Konosuke Furuta | Department of Neurology, Murakami Karindoh Hospital, Fukuoka, Japan |
| Tetsushi Takahashi | Department of Neurology, The Aichi Prefectural Federation of Agricultural Cooperatives for Health and Welfare, Kainan Hospital, Aichi, Japan |
| Hideyuki Matsumoto | Department of Neurology, Mitsui Memorial Hospital, Tokyo, Japan |
| Ohta Masahiko | Department of Neurology, Hyogo Prefectural Amagasaki General Medical Center, Hyogo, Japan |
| Akatsuki Kubota | Department of Neurology, The University of Tokyo, Tokyo, Japan |

#### SECTION S2: PHARMACOKINETICS

1) Ropinirole concentration in blood

Blood samples were drawn at the following time points:

- After interim registration (Week 12)
- After the start of study treatment (Week 2)
- After dose escalation (Week 3−8)
- Before study drug administration at Week 13 and 24
- At the time of discontinuation

For subjects who proceeded to the open-label extension period, blood samples were drawn at similar time points as mentioned above and additional time points as listed below:

(A) Dose at the end of the double-blind period: 2 mg to 4 mg

At Weeks 26, 37, and 48 after the start of study treatment, or at the end of treatment

(B) Dose at the end of the double-blind period: 6 mg

At Weeks 27, 38, and 49 after the start of study treatment, or at the end of treatment

(C) Dose at the end of the double-blind period: 8 mg to 16 mg

At Weeks 28, 39, and 50 after the start of study treatment, or at the end of treatment

2) Ropinirole concentration in cerebrospinal fluid (CSF)

CSF samples were collected using lumbar puncture:

- Before the start of the first dose (3 days before to the day of the first dose)
- At Week 24 after the start of study treatment
- At the time of discontinuation

For subjects who proceeded to the open-label extension period, CSF samples were collected at similar time points as shown above, and additional time points as listed below.

(A) Dose at the end of the double-blind period: 2 mg to 4 mg

At Week 48 after the start of study treatment or at the end of treatment

(B) Dose at the end of the double-blind period: 6 mg

At Week 49 after the start of study treatment or at the end of treatment

(C) Dose at the end of the double-blind period: 8 mg to 16 mg

At Week 50 after the start of study treatment or at the end of treatment

(D) At the time of discontinuation after the final follow-up.

Shimadzu Tecno-Research, Inc. performed the measurements and submitted the results directly to the data management department to maintaining the blind of the study.

#### SECTION S3: SUPPLEMENTARY METHOD

The first draft of the manuscript was written by the first author and the co-authors provided medical writing assistance. K Pharma reviewed the manuscript and provided feedback to authors. Authors had full editorial control of the manuscript and provided their final approval of all content. All authors vouch for the accuracy and completeness of the data, for the fidelity of the trial to the protocol, and for the complete reporting of adverse events. Confidentiality agreements were in place between authors and K Pharma or GlaxoSmithKline (GSK).

Description of the similarity of interventions

GSK provided 2mg and 8mg control-released ropinirole hydrochloride in a light red white or reddish brown, oval and film-coated tablet. The placebo drug was matched to the study drug for taste, color, and size, and contained hypromerose, lactose hydrate, glycerin fatty acid ester, D-mannitol, sodium carmellose, hydrogenated oil, povidone, dextrin, magnesium stearate, light anhydrous silicic acid, yellow iron sesquioxide, titanium oxide, macrogol 400, and iron sesquioxide.

Genetic screening for familial ALS

We performed targeted sequencing by using next generation sequencer and our original ALS screening panel v2 (Total 63 genes: *ALS2, SETX, TUBA4A, PLEKHG5, DNAJB2, ANG, SIGMAR1, CHCHD10, SMN1, DYNC1H1, ATXN2, SOD1, TBK1, TRPV4, BICD2, C9ORF72, SPG11, EWSR1, VRK1, FBXO38, CHMP2B, TAF15, SQSTM1, HSPB1, ASAH1, DAO, TARDBP, SS18L1, HSPB3, EXOSC8, DCTN1, UBQLN2, GLE1, HSPB8, EXOSC3, FIG4, VAPB, GRN, ATL1, SLC52A3, FUS, VCP, ZNF512B, SPAST, SLC52A2, NEFH, ERBB4, ATP7A, AARS, HEXB, OPTN, HNRNPA1, BSCL2, REEP1, MAPT, PFN1, HNRNPA2B1, GARS, SLC5A7, PRPH, MATR3, IGHMBP2, UBA1*) and exome analysis to examine mutations related to ALS (Neurobiol Aging 2017;53:194.e1-194.e8.).

Post-hoc analyses

1) Ratio of change in the ALSFRS-R score every 4 weeks between the 12-week double-blind period (the latter of 12 weeks) and 24-week open label extension period assessments

The change in the ALSFRS-R score every 4 weeks during the double-blind period (the latter of 12 weeks) and the change in the ALSFRS-R score every 4 weeks during the 24-week open label extension period were calculated to obtain the ΔALSFRS-R ratio by the following formula.

$$\Delta\text{ALSFRS-R ratio} = \frac{\text{Change in ALSFRS-R score every 4 weeks during the 24-week open label extension period}}{\text{Change in ALSFRS-R score every 4 weeks during the 12-week double-blind period (the latter of 12 weeks)}}$$

The change in the ALSFRS-R score every 4 weeks during t the 12-week double-blind period (the latter of 12 weeks) and during t the 24-week open label extension period were calculated based on the slope of a regression equation built by using the ALSFRS-R score measured for each subject as a response variable and the number of days from the start day of treatment at each measurement time point as an explanatory variable.

The analysis was performed on the FAS. Summary statistics (mean, SD, minimum, median, maximum, and 95%CI) were calculated for the measured value and the change from baseline in the ΔALSFRS-R ratio during the 12-week double-blind period (the latter of 12 weeks) and during the 24-week open label extension period by treatment group. To confirm whether there was any significant change from the 12-week double-blind period (the latter of 12 weeks) to the 24-week open label extension period, a null hypothesis that the mean change from baseline at each time point was 0 was tested by treatment group using a one-sample t-test. In addition, to confirm whether there was any difference in the change between the groups, the least squares mean difference and the two-sided 95%CI were calculated using contrasts by an ANCOVA model with baseline value as a covariate. The baseline value was defined as the value on the day of the first dose of study treatment. If a significant deviation was found in the distribution of the dependent variable, a non-parametric approach was used as needed.

2) Composite endpoint as a sum of Z-transformed scores at 39 and 50 weeks

The analysis was performed on the FAS. Z-transformed scores at each time point were calculated using values of each item at Weeks 39 and 50 of the open label extension period and summed as the composite endpoint. Summary statistics (mean, SD, minimum, median, maximum, and 95%CI) were calculated for the composite endpoint at each time point and the change in the composite endpoint from 12 weeks after interim registration by treatment group. To confirm whether there was any difference in the change in the composite endpoints between the groups, MMRM analysis, with change from 12 weeks after interim registration as a dependent variable, was performed. In addition, summary statistics were calculated for the composite endpoint of Z-transformed scores obtained at Weeks 39 and 50 of the open label extension period. For differences between two groups, MMRM analysis was performed with the composite endpoint calculated from the change as a dependent variable. If a significant deviation was found in the distribution of the dependent variable, a non-parametric approach was used as needed.

3) The following items in the composite endpoint as a sum of Z-transformed scores

•ALSFRS-R sub-score of each domain (bulbar function, limb function, and respiratory function)

•Simple respiratory function test (FEV1, FEV6)

•Detailed respiratory function test (%FVC, FEV1%)

•MMT score (neck flexion, elbow flexion [right], elbow flexion [left], wrist extension [right], wrist extension [left], hip flexion [right], hip flexion [left], ankle dorsiflexion [right], ankle dorsiflexion [left])
•Quantitative muscle strength (neck flexion, elbow flexion [right], elbow flexion [left], wrist extension [right], wrist extension [left], hip flexion [right], hip flexion [left], ankle dorsiflexion [right], ankle dorsiflexion [left])
•Grip strength (right, left)
•Pinch strength (right, left)
•Modified Norris Scale (bulbar symptom score)
•Tongue pressure
•Body weight
•ALSAQ-40 score (physical mobility, ADL/independence, eating and drinking, communication, emotional functioning)
•Creatinine

The analysis was performed on the FAS. Summary statistics (mean, SD, minimum, median, maximum, and 95%CI) of all items were calculated for the double-blind period, open label extension period, and overall treatment period by treatment group. To confirm whether there was any significant change in each group, a null hypothesis that the change from baseline at each time point was 0 was tested by treatment group using a one-sample t-test. In addition, to confirm whether there was any difference in the change between the groups, MMRM analysis, with the change from baseline as a dependent variable, was performed for the double-blind period, open label extension period and overall treatment period. Baseline was defined as the day of the first dose of study treatment. If a significant deviation was found in the distribution of the dependent variable, a non-parametric approach was used if necessary.
