## Supplementary material for "Ropinirole hydrochloride for amyotrophic lateral sclerosis: A single-center, randomized feasibility, double-blind, placebo-controlled trial": SECTION S4-5

### SECTION S4: SUPPLEMENTARY FIGURES

Figure S1. Ropinirole Hydrochloride Remedy for Amyotrophic Lateral Sclerosis (ROPALS) Study Design.

Trial participants who completed the run-in period were randomized to receive either ropinirole or placebo at a 3:1 ratio. Due to dynamic allocation and allocation adjustment factors, 13 participants were assigned to receive ropinirole and 7 participants were assigned to receive placebo (approximately a 2:1 ratio). The in vitro evaluation of drug effects and exploration of new biomarkers using patient iPSC-derived motor neurons are ongoing.

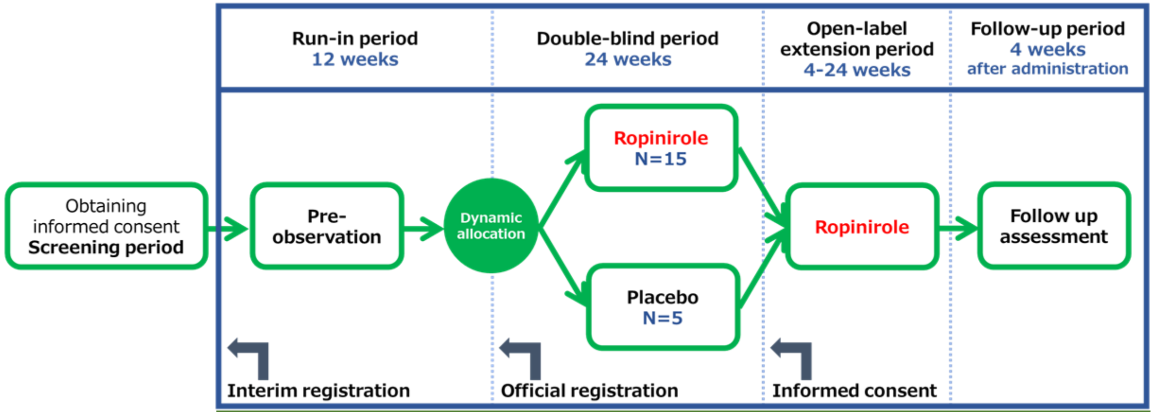


| STUDY ENDPOINTS | |
|---|---|
| **Primary** | Type, frequency, and severity of adverse events (AEs), and time course of laboratory test values, and intergroup difference in the proportion of discontinued subjects during the 24-week double-blind period |
| **Secondary** | 1) Ratio of change in the ALSFRS-R score every 4 weeks between pre- and post-treatment assessments<br>2) ΔALSFRS-R score during the double-blind period<br>3) ΔALSFRS-R score during the open-label extension and the entire period<br>4) CAFS score during the double-blind, the open-label extension, and the entire period<br>5) Composite endpoint as a sum of Z-transformed scores<br>6) Time to death or a specified state of disease progression<br>7) Time to %FVC of ≤50%<br>8) Time to decrease of at least 6 points in the ALSFRS-R score<br>9) Amount of physical activity before and after treatment during the double-blind period<br>10) Proportion of discontinued subjects during the period from Day 1 of the double-blind period to the final assessment of the open-label extension period<br>11) Other assessment items (ALS severity classification) |
| **Exploratory** | 1) Comparison of the in vitro drug effect evaluation and clinical effect using patients' iPSC-derived motor neurons<br>2) Exploration of new biomarkers for diagnosis, pathology, and drug effect evaluation<br>3) Search of known FALS-related genes<br>4) Zarit Caregiver Burden Interview<br>5) Ropinirole concentrations in blood and spinal fluid<br>6) Objective muscle mass determined using computed tomography (CT) for skeletal muscle |

Figure S2. Measured Concentration of Ropinirole in Plasma and CSF over The Administration Period.

The figure shows the mean (95%CI) concentration of ropinirole in plasma and CSF in all trial participants in the double-blind period.

Abbreviation: CSF = Cerebrospinal fluid

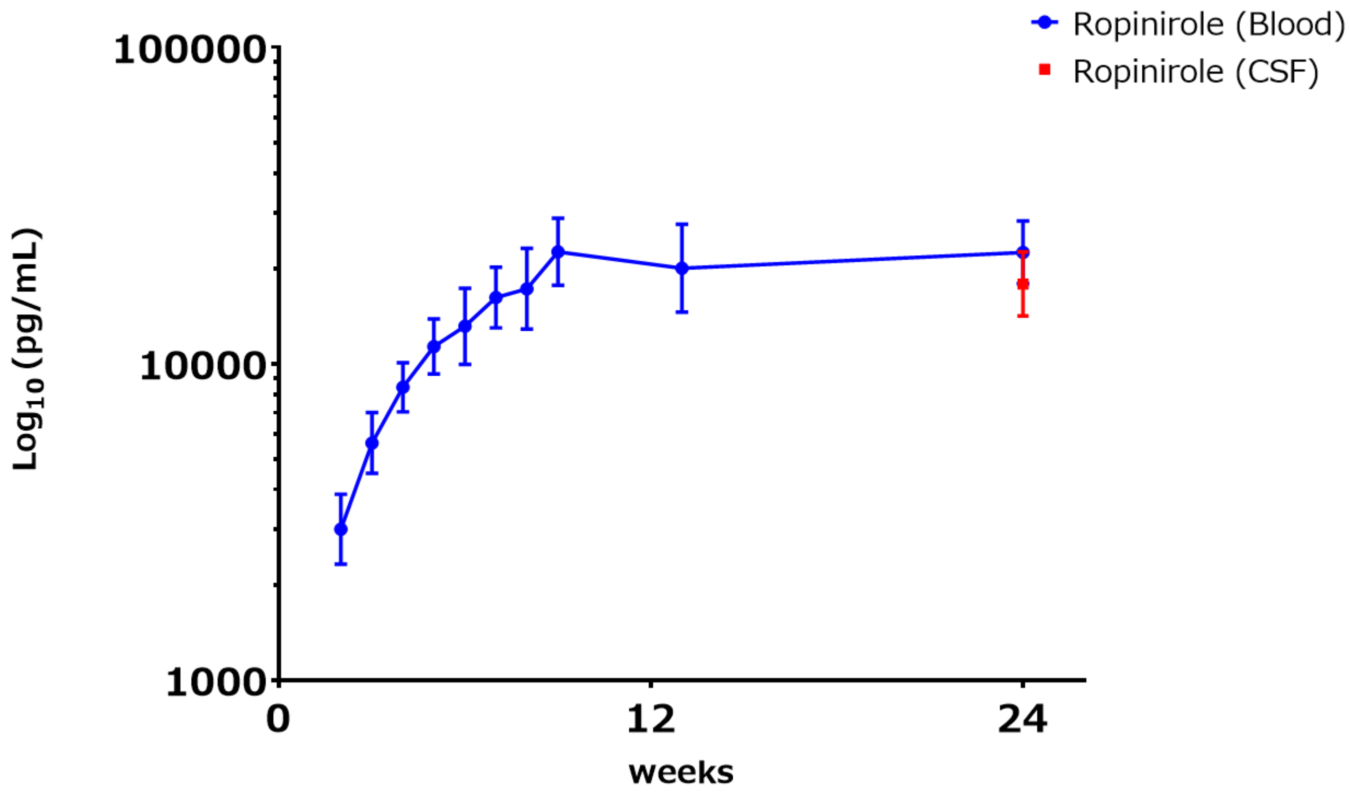


| Samples | Plasma | | | | | | | | | | | CSF | |
|---|---|---|---|---|---|---|---|---|---|---|---|---|---|
| Requip CR (mg) | 0 | 2 | 4 | 6 | 8 | 10 | 12 | 14 | 16 | 16 | 16 | 0 | 16 |
| No. of Participants | 18 | 18 | 18 | 18 | 18 | 18 | 18 | 18 | 18 | 17 | 14 | 13 | 13 |

Figure S3. Effect of Ropinirole Treatment on ALSFRS-R Each Score.

Estimated change from baseline in ALSFRS-R each score every four weeks (FAS, entire period).

‡ Differences >0 indicate a positive treatment effect of ropinirole.

I bars represent the 95%CI.

Abbreviations: ALSFRS-R = Revised ALS Functional Rating Scale, FAS = Full Analysis Set

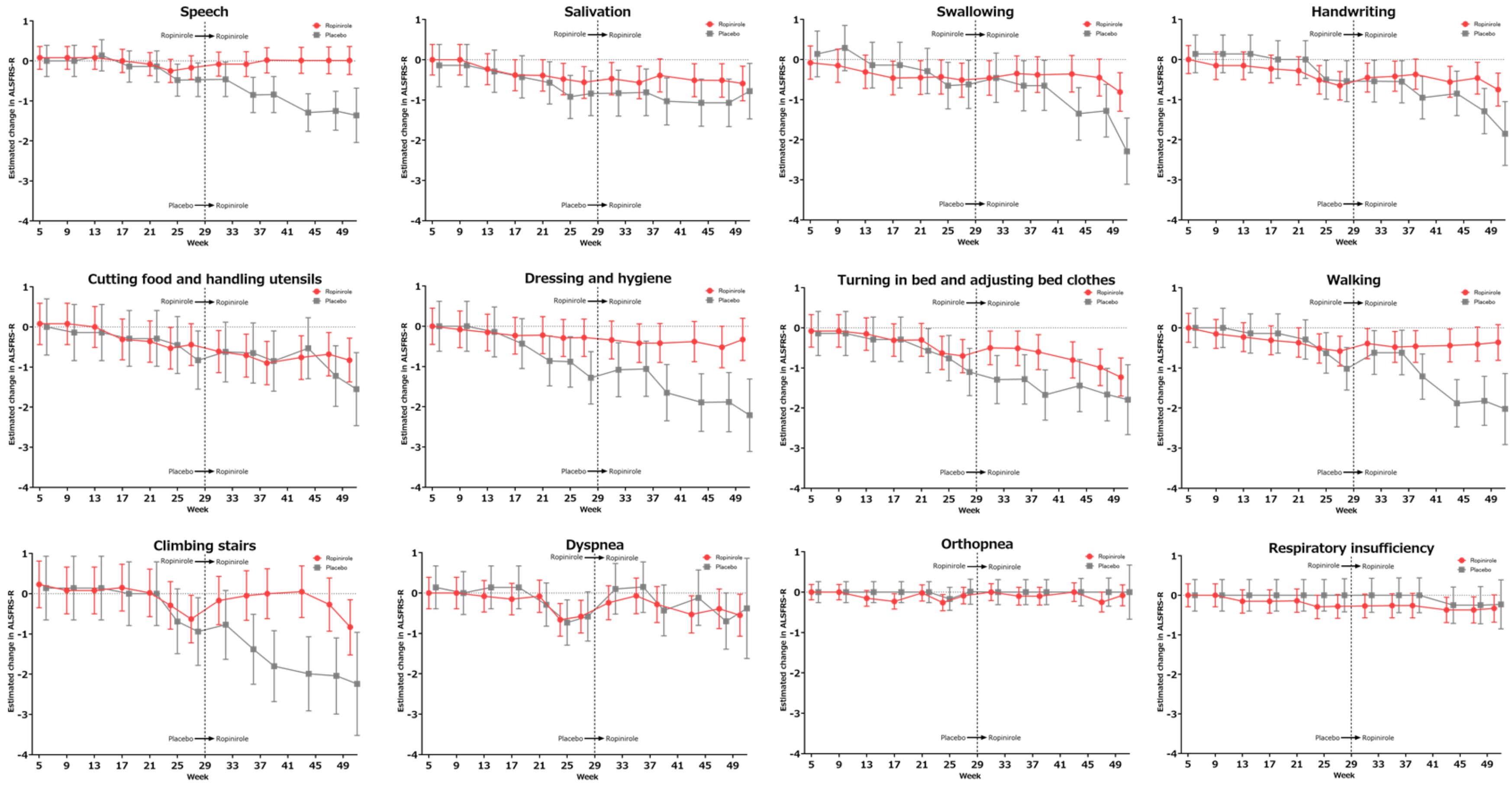

Figure S4. Changes in Four Subdomains of the ALSFRS-R Score (z-score) from baseline over 48 Weeks.

The upper and lower limb domains were combined and presented as “motor function”.

Lower scores indicate worse function. I bars represent the SE.

‡ Difference >0 indicates a positive treatment effect of ropinirole.

Abbreviations: ALSFRS-R = Revised ALS Functional Rating Scale, SE = Standard Error.

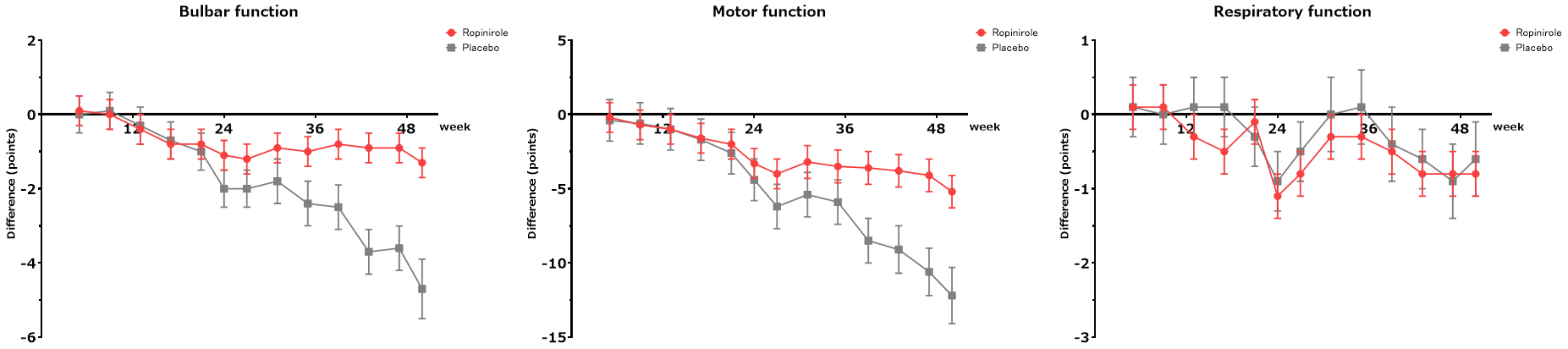

Figure S5. Changes in Muscle Strength, Grip Strength, and Pinch Strength (z-score) from Baseline over 48 Weeks.

Lower scores indicate worse function. I bars represent the SE.

‡ Difference >0 indicates a positive treatment effect of ropinirole.

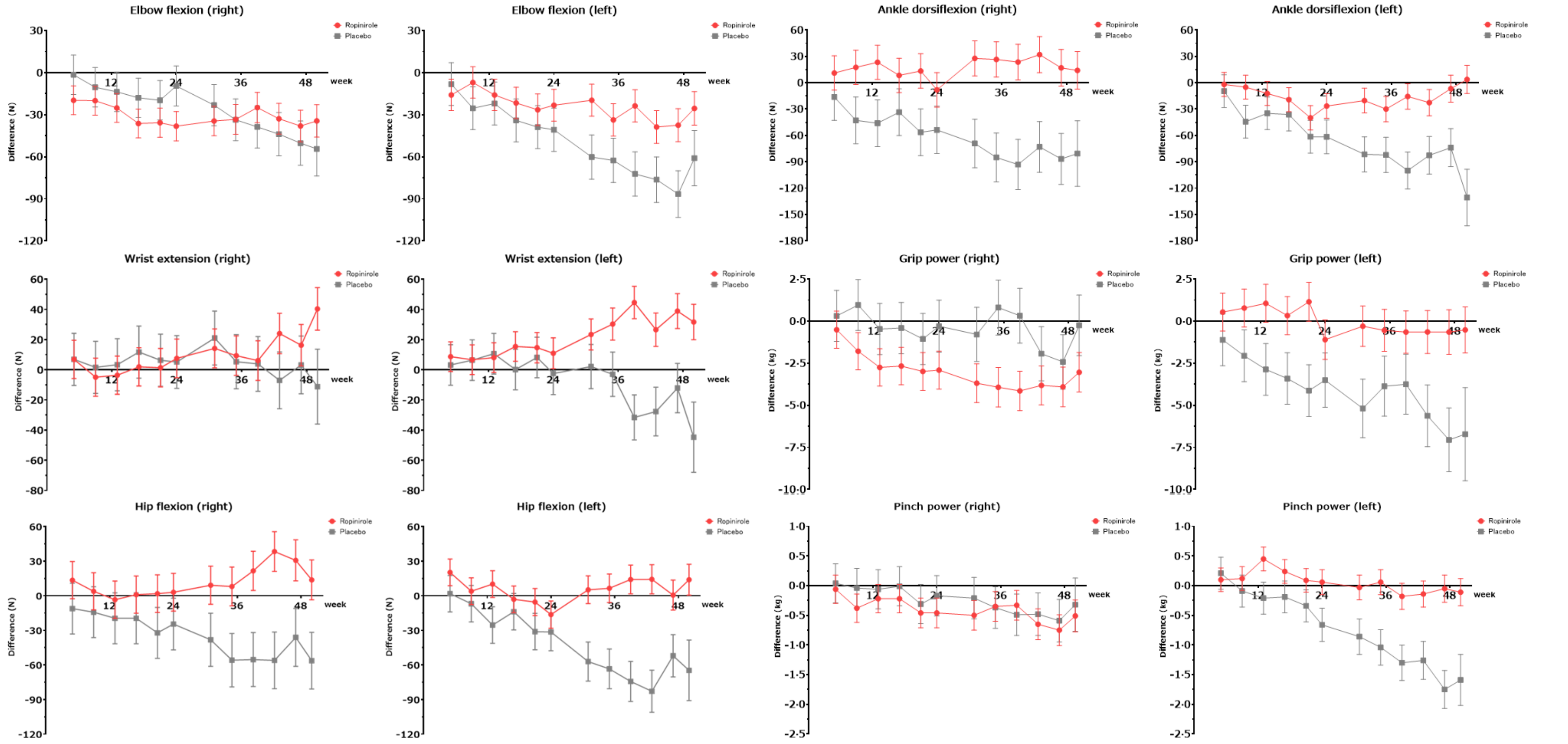

Figure S6. Changes in %FVC, Norris bulbar scale, tongue pressure, body weight and serum creatinine (z-score) from Baseline over 48 Weeks. Lower scores indicate worse function. I bars represent the SE.

‡ Difference >0 indicates a positive treatment effect of ropinirole.

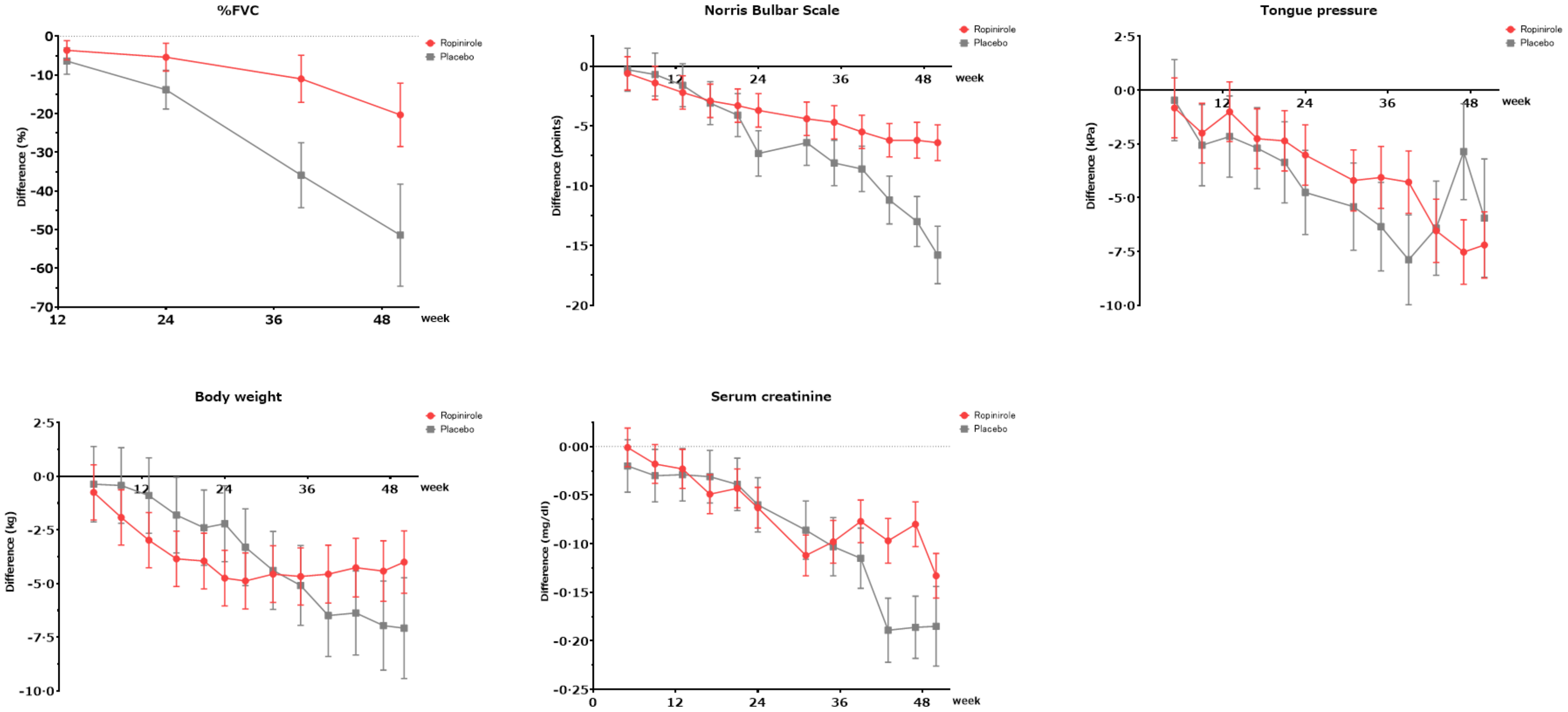

Figure S7. Changes in ALSAQ40 Scores (z-score) from Baseline over 48 Weeks.

Higher scores indicate worse function. I bars represent SE.

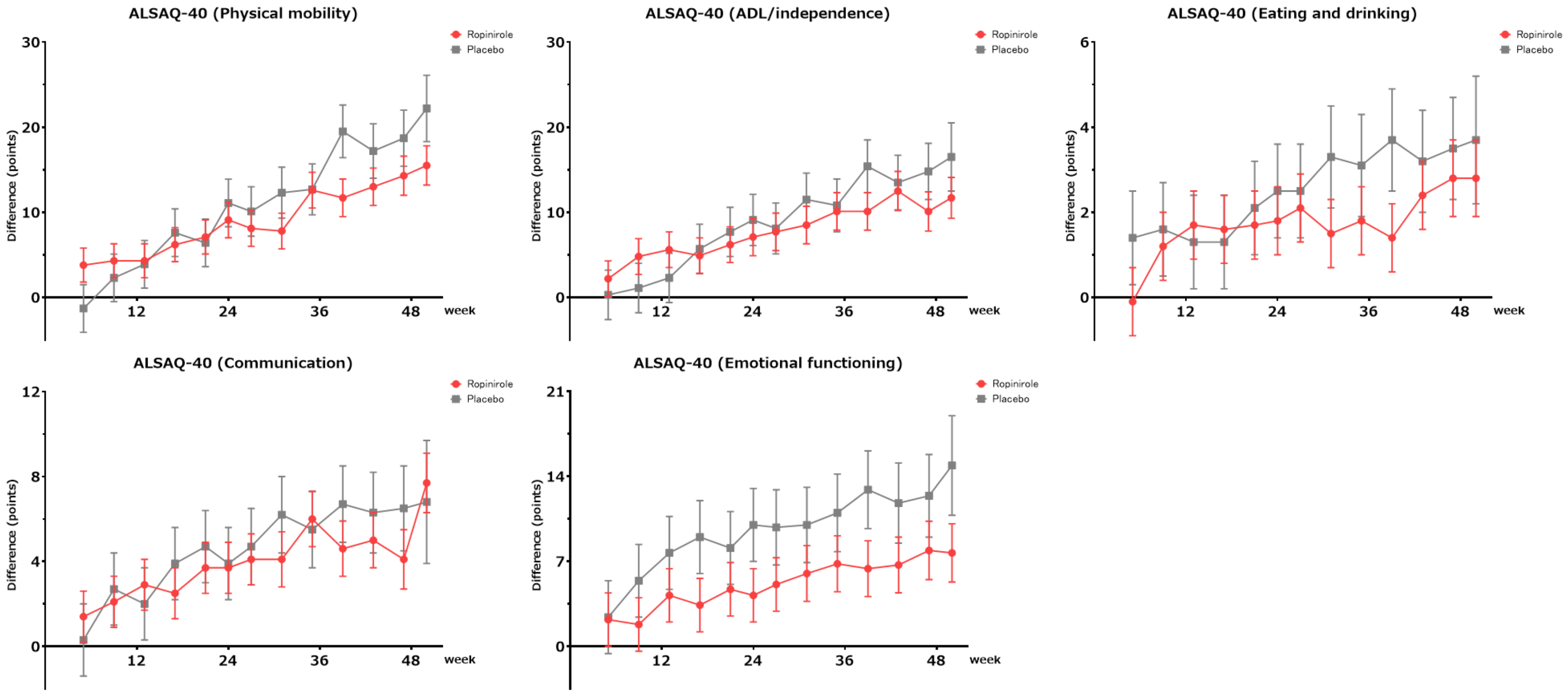

Figure S8. Japanese ALS Severity Scale Categories of Participants at The End of 24 Weeks and 48 Weeks.

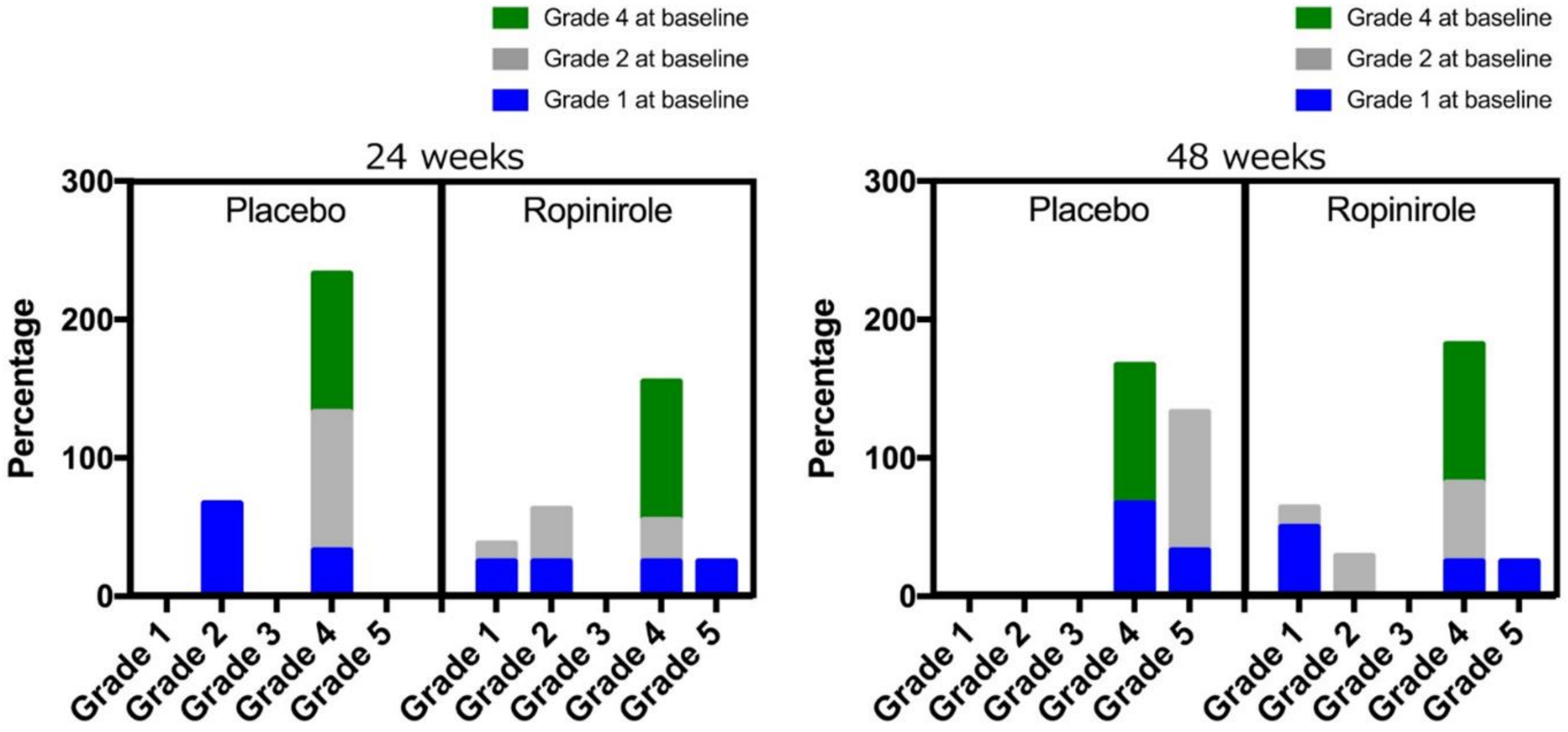

Figure S9. Effect of Ropinirole on Ferritin, 8-OHdG, and HS-CRP Concentrations over 48 Weeks.

The mechanism of action for ropinirole in ALS is unknown; we speculate that it could be D2 receptor-dependent or independent. Biomarkers analyses of ferritin, 8-hydroxy-2'-deoxyguanosine (8-OHdG), high-sensitivity C-reactive protein (HS-CRP), neurofilament, and lipid peroxidation were performed.

The Student's t-test was used for intergroup comparisons at each time point. The 48–50-week dataset included the discontinued period (44 and 48 weeks). Outlier (1) was excluded using the ROUT method. I bars represent the SE.

Abbreviations: 8-OHdG/Cre = 8-hydroxy-2'-deoxyguanosine/creatinine, HS-CRP = high-sensitivity C-reactive protein, CSF = cerebrospinal fluid, NF-L = Neurofilament, MDA = Malondialdehyde.

(A) Effect of ropinirole on Ferritin, 8-OHdG, and HS-CRP Concentrations over 48 Weeks.

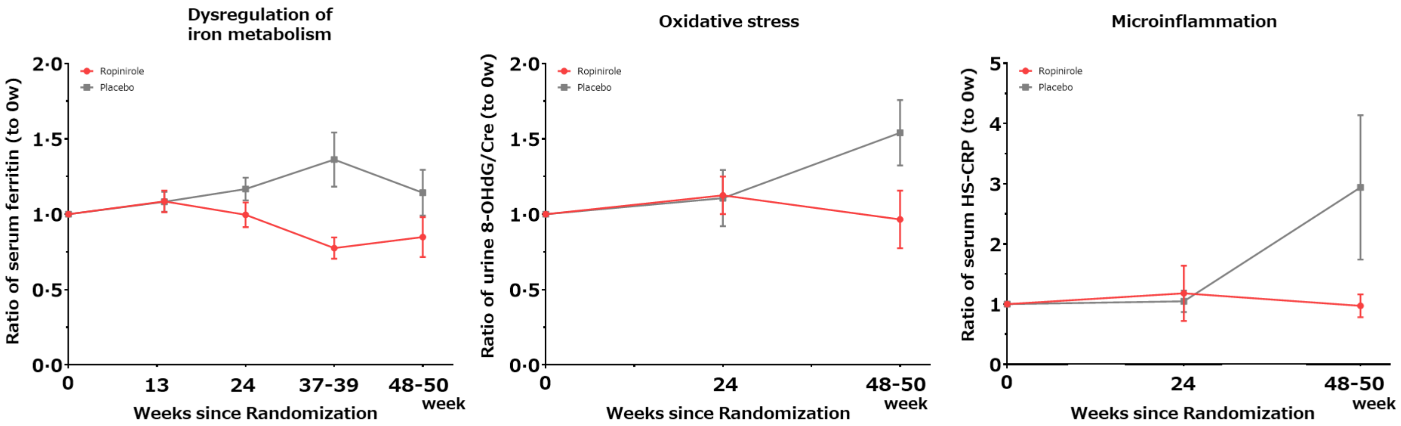


The change value of the proteins to the baseline

| Time points | | 13w | 24w | 37-39w | 48-50w (or the end) |
|---|---|---|---|---|---|
| **No. of participants (Ropinirole/Placebo)** | | 13/7 | 12/6 | 10/5 | 9/3 |
| **Ratio of serum ferritin** | **Difference (95%CI) P value** | 0·00 (-0·23 to 0·24) 0·979 | -0·17 (-0·44 to 0·10) 0·202 | -0·59 (-0·93 to -0·25) 0·003 | -0·29 (-0·85 to 0·26) 0·266 |
| **Ratio of Urine 8-OHdG/Cre** | | NA | 0·02 (-0·45 to 0·48) 0.934 | NA | -0·57 (-1·38 to 0·23) 0.142 |
| **Ratio of HS-CRP** | | NA | 0·13 (-1·29 to 1·55) 0·844 | NA | -1·97 (-3·54 - -0·39) 0·019 |

(B) Effect of Ropinirole on Plasma and CSF Neurofilament Concentrations over 48 Weeks.

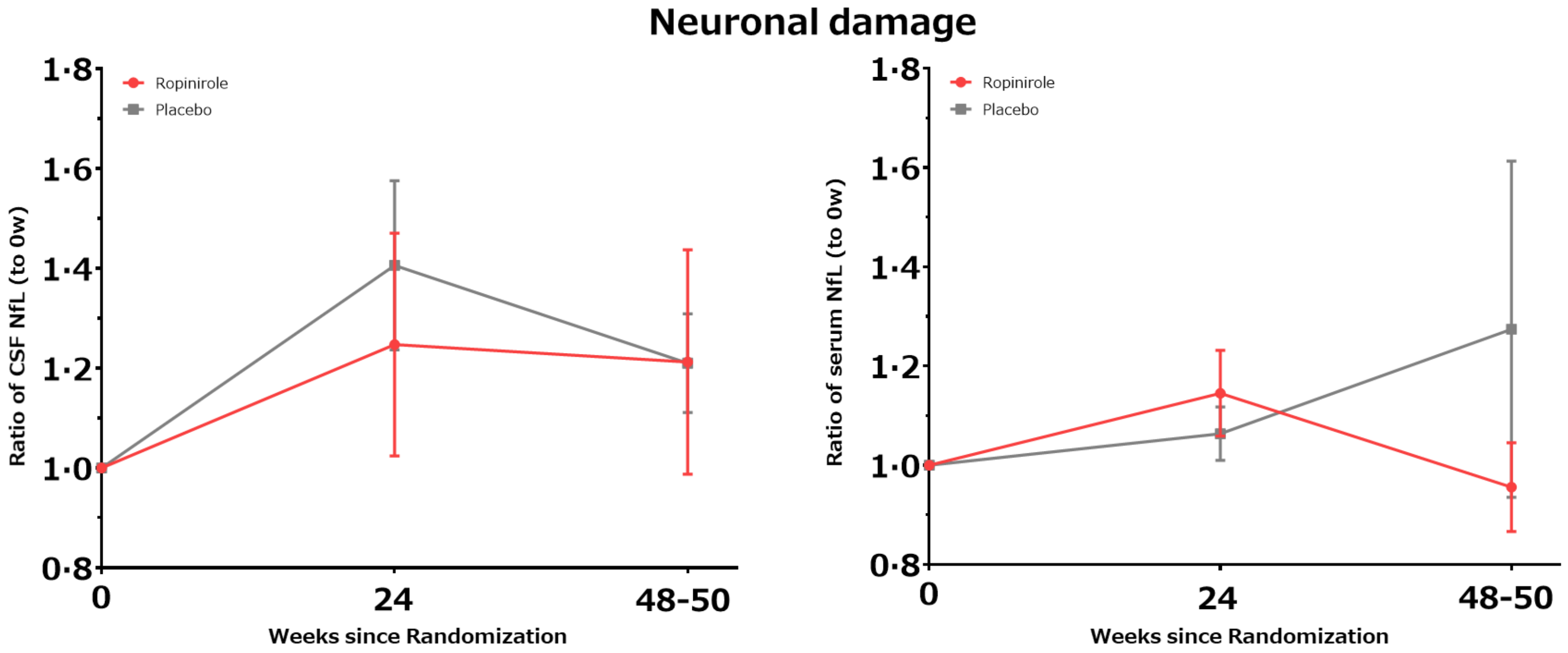


The change value of the NfL to the baseline

| | | CSF | | Serum | |
|---|---|---|---|---|---|
| **Time points** | | 24w | 48-50w (or the end) | 24w | 48-50w (or the end) |
| **No. of participants (Ropinirole/Placebo)** | | 11/6 | 4/2 | 12/6 | 9/3 |
| **Ratio of NfL** | **Difference (95%CI) P value** | -0·16 (-0·86 to 0·55) 0.638 | 0·00 (-0·95 to -0·95) 0.995 | 0·08 (-0·19 to 0·36) 0.539 | -0·32 (-0·86 to 0·23) 0.219 |

(C) Effect of Ropinirole on Lipid Peroxidation over 48 Weeks. Lipid peroxidation was measured as the level of MDA determined by the thiobarbituric acid reaction following the manufacturer's instructions (TBARS Assay Kit, Cayman). The absorbance was measured at 540 nm. The concentration of MDA was expressed as μM of MDA per mg of protein using an iMark Microplate Absorbance Reader.

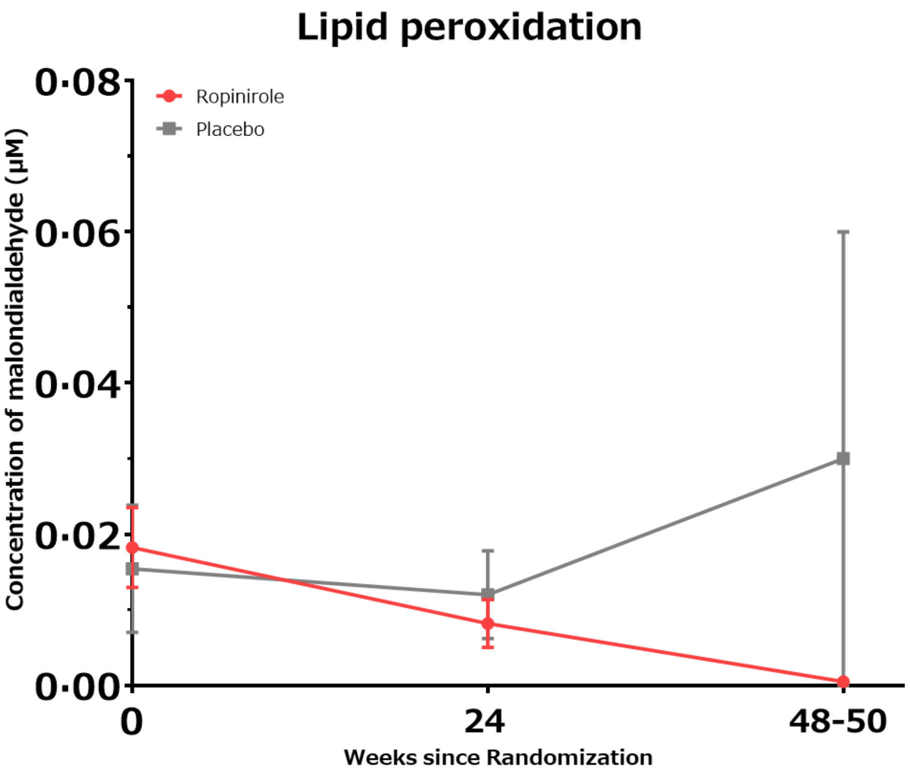


The change value of the malondialdehyde to the baseline

| | | CSF | | |
|---|---|---|---|---|
| **Time points** | | 0w | 24w | 48-50w (or the end) |
| **No. of participants (Ropinirole/Placebo)** | | 13/7 | 11/6 | 4/2 |
| **Ratio of MDA** | **Difference (95%CI) P value** | 0·003 (-0·017 to 0·023) 0.770 | 0·006 (-0·017 to 0·009) 0.537 | -0·030 (-0·081 to 0·022) 0.184 |

Figure S10. Effect of pretreatment of ropinirole hydrochloride for low concentration on ALS phenotype.
(A) Relative quantitative data for neurite length, LDH leakage, FUS aggregation, and SGs in LMNs derived from FUS-mutant iPSCs.
(B) Relative quantitative data for neurite length, LDH leakage, phosphorylated TDP-43 (pTDP-43) aggregation, and CC3-positive neurons in LMNs derived from TDP-43-mutant iPSCs.
(n = 3 independent experiments; mean ± standard deviation.; *$p<0.05$, **$p<0.01$; one-way ANOVA followed by Dunnett's multiple comparisons test)
(C) Efficacy evaluation of low dose ropinirole hydrochloride more than 0.1 nM for ALS phenotypes; shorter neurite length, leaked LDH, increased SG formation and increased FUS aggregates in FUS-mutant LMNs, and shorter neurite length, leaked LDH, increased pTDP-43 aggregates and increased CC3-positive neurons.
Abbreviation: ROPI: Ropinirole hydrochloride, ALS: Amyotrophic lateral sclerosis, LDH; Lactate dehydrogenase, FUS; Fused in Sarcoma, SG; Stress granule, LMN; Lower motor neurons, TDP-43; TAR DNA-binding Protein of 43kDa, CC3; cleaved caspase 3.
The detailed methods are written in the previous paper (Fujimori K, et al. Nat Med 2018).

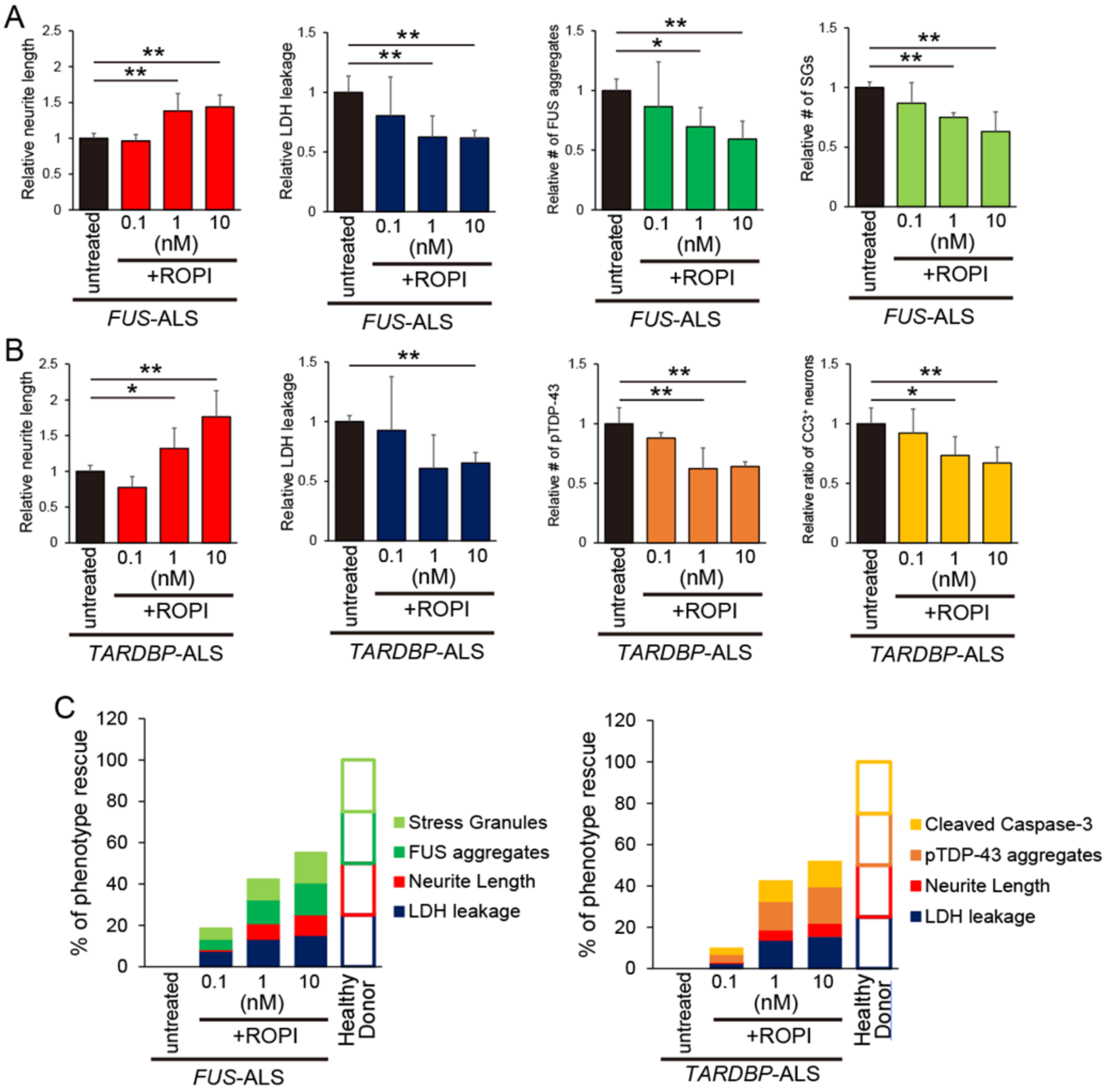

A
Relative neurite length
Relative LDH leakage
Relative # of FUS aggregates
Relative # of SGs
untreated
0.1
1
10
(nM)
+ROPI
FUS-ALS
B
Relative # of pTDP-43
Relative ratio of CC3+ neurons
TARDBP-ALS
C
% of phenotype rescue
Stress Granules
FUS aggregates
Neurite Length
LDH leakage
Healthy Donor
Cleaved Caspase-3
pTDP-43 aggregates

Figure S11. The expression of dopamine D2 receptor (D2R) in anterior horn cells of human spinal cord.

We used an anti-dopamine D2R antibody (NLS1403, Novus Biologicals, CO, USA) and formalin-fixed, paraffin-embedded sections of a healthy human thoracic spinal cord. Upper figures are low magnification and lower figures are enlarged images within red squares in the upper figures.

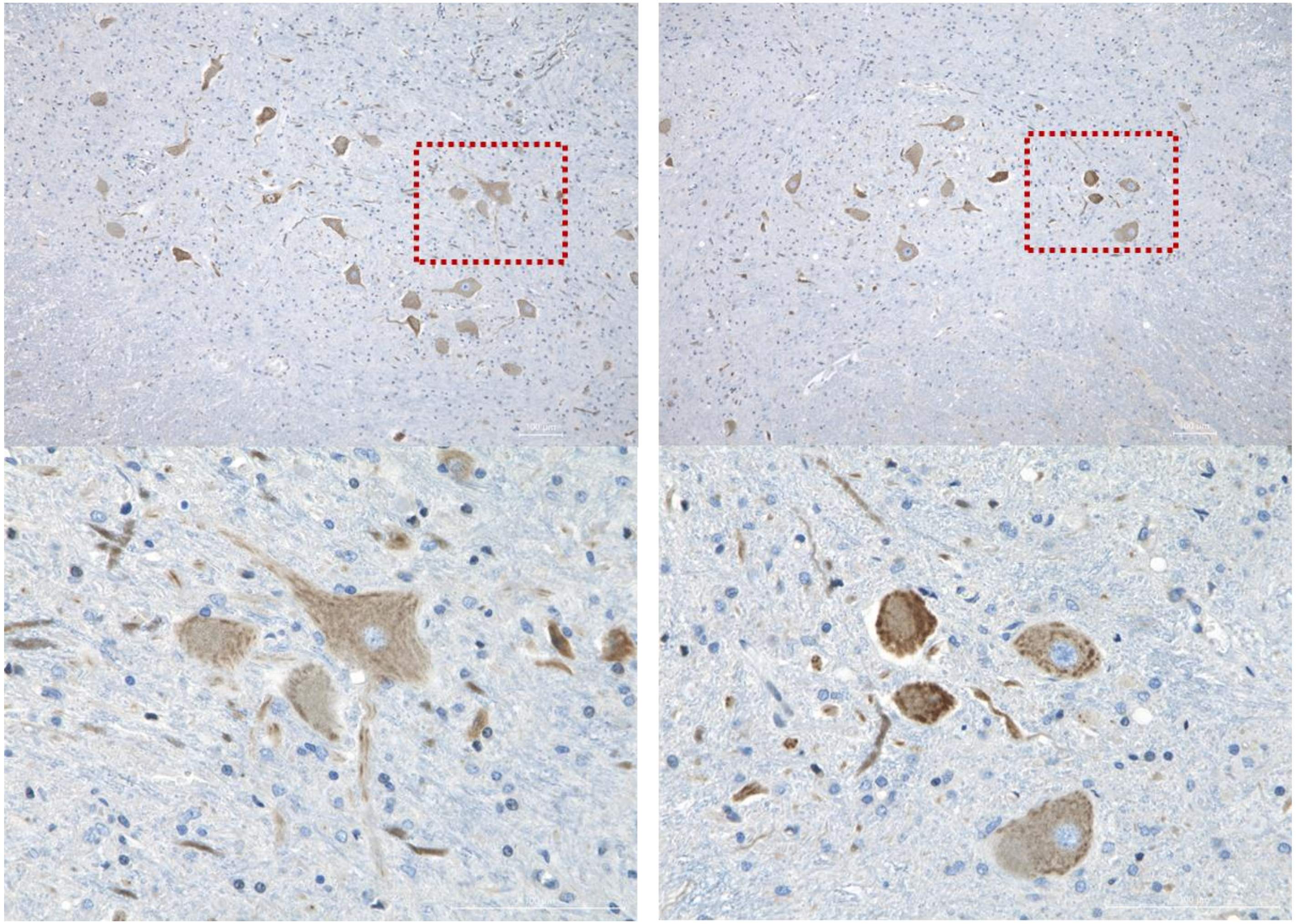

Figure S12. The expression of D2 receptor (D2R) in human iPSC-derived lower motor neurons.

We induced lower motor neurons from healthy-derived iPSCs (201B7) by a modified protocol in the previous report (Goto K, et al. Mol Ther Methods Clin Dev 2017) and stained them by using an anti-dopamine D2R antibody (clone 3D9, Merck Millipore, MA, USA), anti-β3-tubulin antibody (T8660, Sigma-Aldrich, MO, USA) and Hoechst33342 for nuclear staining.

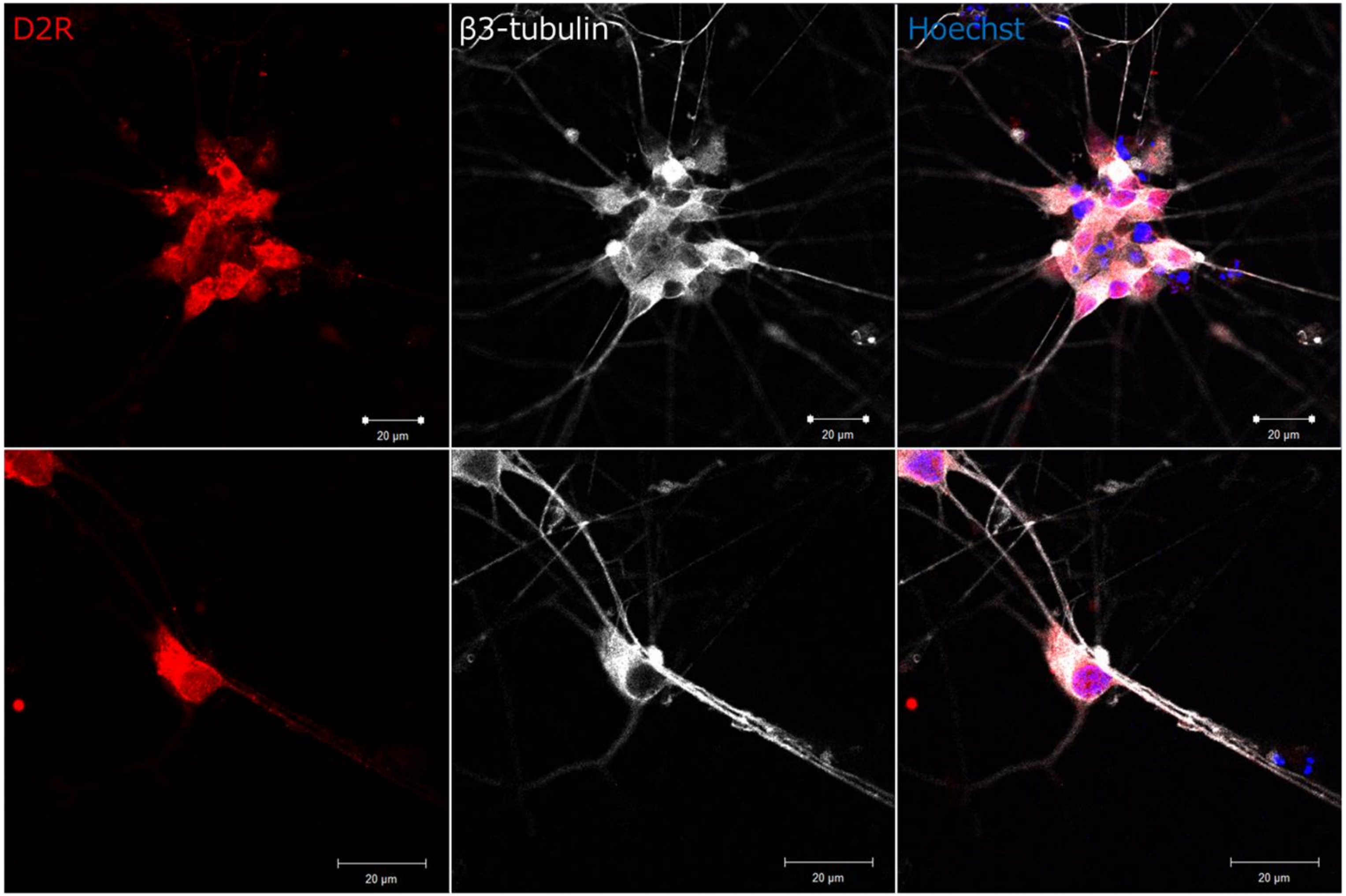

Figure S13. The Transition of Estimated Change in ALSFRS-R during Entire Period for RR-bulbar-, RR-nonbulbar-, PR-bulbar-, and PR-nonbulbar-Onset Groups.

(A) The transition of estimated change in ALSFRS-R of RR-group), and (B) The transition of estimated change in ALSFRS-R of PR-group. There was no significant difference in the estimated change in ALSFRS-R between the bulbar-onset and nonbulbar-onset groups for either ropinirole or placebo at any time point.

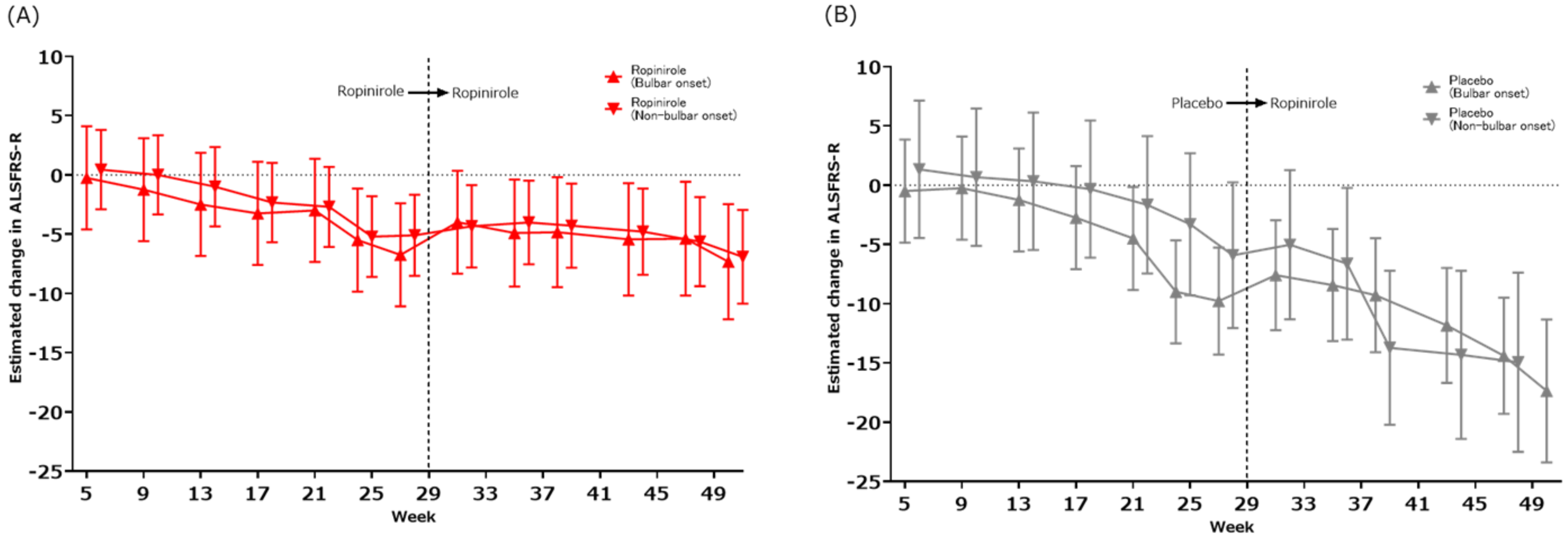

### SECTION S5: SUPPLEMENTARY TABLES

Table S1. Plasma Concentration of Ropinirole When Drug Dose Was Increased to A Maximum of 16 mg.

The table shows the mean (standard deviation [SD]) concentration of ropinirole in plasma and CSF during the double-blind period.

| Ropinirole | Plasma concentration (pg/mL) | | | Case |
|---|---|---|---|---|
| mg | Geometric mean | 95% CI | CV (%) | N |
| 0 | 0 | | | 18 |
| 2 | 3004 | 2330 - 3874 | 59·56 | 18 |
| 4 | 5623 | 4508 - 7015 | 49·64 | 18 |
| 6 | 8433 | 7052 - 10084 | 38·33 | 18 |
| 8 | 11347 | 9291 - 13859 | 42·17 | 18 |
| 10 | 13150 | 9971 - 17343 | 44·33 | 18 |
| 12 | 16199 | 12991 - 20198 | 46·78 | 18 |
| 14 | 17266 | 12875 - 23153 | 54·20 | 18 |
| 16 | 22593 | 17705 - 28830 | 43·46 | 18 |
| 16 | 20058 | 14573 - 27608 | 43·30 | 17 |
| 16 | 22510 | 17923 - 28271 | 39·30 | 14 |
| **Ropinirole** | **CSF concentration (pg/mL)** | | | **Case** |
| **mg** | **Geometric mean** | **95% CI** | **CV (%)** | **N** |
| 0 | 0 | | | 13 |
| 16 | 17884 | 14186-22547 | 38·25 | 13 |

Table S2. LS mean Change from Baseline in ALSFRS-R Score (Double-blind and Open-label extension period, FAS)

Abbreviations: LS = least-squares, DB = Double-blind, OL = Open-label extension

| | **Least-Squares Mean Change per 4 weeks** | | **Least-Squares Difference** |
|---|---|---|---|
| | **DB period (24 weeks)/Run-in period (12 weeks)** | | |
| | Ropinirole hydrochloride (N=13) | Placebo (N=7) | |
| Ratio of change in ALSFRS-R score (95% CI) | 1·042 (0·249 to 1·835) | 1·254 (0·165 to 2·344) | -0·212 (-1·577 to 1·153) |
| | **OL period (24 weeks)/DB period (the latter of 12 weeks)** | | |
| | Ropinirole hydrochloride (N=11) | Placebo (N=5) | |
| Ratio of change in ALSFRS-R score (95% CI) | 0·147 (-1·052 to 1·346) | 1·939 (0·159 to 3·719) | -1·792 (-3·941 - 0·357) |

Table S3. Estimate Rate of Decline in ALSFRS-R Score and Level of Activity over 48 weeks.

| | Mean Change per Month | | | | | | | |
|---|---|---|---|---|---|---|---|---|
| | Ropinirole hydrochloride | Placebo | Difference‡(95%CI) | P value | Ropinirole hydrochloride | Placebo | Difference‡(95%CI) | P value |
| | ALSFRS-R (Prediction value) | | | | Amount of activity (Prediction value) | | | |
| Double-blind period | | | | | | | | |
| 1-4w | 0·23±0·50 | 0·29±0·68 | -0·05 (-1·82 - 1·71) | 0.949 | 27·9±59·9 | -23·9±81·6 | 51·8 (-160·9 – 264·4) | 0.615 |
| 5-8w | -0·38±0·58 | 0·14±0·80 | -0·53 (-2·60 - 1·55) | 0.600 | 86·2±89·7 | 1·0±122·2 | 85·2 (-233·2 – 403·6) | 0.581 |
| 9-12w | -1·46±0·83 | -0·57±1·13 | -0·89 (-3·84 - 2·06) | 0.533 | 108·4±85·4 | -69·9±116·3 | 178·4 (-124·8 – 481·5) | 0.232 |
| 13-16w | -2·62±1·28 | -1·71±1·74 | -0·90 (-5·44 - 3·64) | 0.682 | -9·0±77·6 | -92·7±105·5 | 83·7 (-191·5 – 358·9) | 0.531 |
| 17-20w | -2·90±1·37 | -3·29±1·86 | 0·38 (-4·46 - 5·23) | 0.870 | -16·2±79·2 | -184·4±107·6 | 168·2 (-112·4 – 448·9) | 0.224 |
| 21-24w | -6·16±2·07 | -6·54±2·82 | 0·38 (-6·97 - 7·72) | 0.915 | 67·9±70·9 | -249·6±96·3 | 317·5 (66·3 – 568·7) | 0.016 |
| Open-label extension period | | | | | | | | |
| 29-32w | 1·36±1·17 | 1·60±1·73 | -0·24 (-4·43 - 3·96) | 0.910 | -63·6±103·3 | 71·5±153·2 | -135·1 (-531·4 – 261·3) | 0.477 |
| 33-36w | 1·18±1·18 | 0·40±1·73 | 0·78 (-3·43 - 4·98) | 0.712 | 30·4±85·5 | 298·5±122·6 | -268·2 (-588·8 – 52·4) | 0.094 |
| 37-40w | 0·89±1·19 | -3·00±1·73 | 3·89 (-0·32 - 8·11) | 0.070 | -51·3±119·8 | 33·4±169·7 | -84·6 (-530·1 – 360·8) | 0.690 |
| 41-44w | 0·26±1·21 | -5·12±1·78 | 5·38 (1·07 - 9·70) | 0.015 | 1·9±101·9 | 176·0±150·2 | -174·1 (-563·3 – 215·1) | 0.354 |
| 45-48w | -0·31±1·25 | -7·21±1·81 | 6·89 (2·47 - 11·31) | 0.003 | -84·7±90·8 | 33·0±130·5 | -117·7 (-458·8 – 223·3) | 0.471 |

| | | | | | | | | |
|---|---|---|---|---|---|---|---|---|
| 49-51w (or the end) | -1·94±1·30 | -9·69±2·38 | 7·74 (2·30 - 13·18) | 0.006 | -134·5±102·2 | -369·9±179·2 | 235·4 (-207·0 – 677·9) | 0.273 |
| Entire period | | | | | | | | |
| 1-4w | 0·23±1·36 | 0·29±1·86 | -0·05 (-4·61 - 4·50) | 0.981 | 27·9±91·1 | -23·9±124·1 | 51·8 (-252·1 – 355·7) | 0.737 |
| 5-8w | -0·38±1·36 | 0·14±1·86 | -0·53 (-5·08 - 4·02) | 0.819 | 86·2±91·1 | 1·0±124·1 | 85·2 (-218·7 – 389·1) | 0.581 |
| 9-12w | -1·46±1·36 | -0·57±1·86 | -0·89 (-5·44 - 3·66) | 0.700 | 108·4±91·1 | -69·9±124·1 | 178·4 (-125·5 – 482·3) | 0.248 |
| 13-16w | -2·62±1·36 | -1·71±1·86 | -0·90 (-5·45 - 3·65) | 0.696 | -11·3±91·8 | -92·7±124·1 | 81·3 (-223·5 – 386·1) | 0.599 |
| 17-20w | -2·80±1·37 | -3·29±1·86 | 0·49 (-4·07 - 5·05) | 0.833 | -8·1±92·3 | -184·4±124·1 | 176·3 (-129·0 – 481·7) | 0.256 |
| 21-24w | -5·36±1·38 | -6·82±1·88 | 1·46 (-3·15 - 6·07) | 0.533 | 33·9±92·9 | -219·4±127·5 | 253·3 (-58·1 – 564·7) | 0.110 |
| 25-28w | -5·75±1·39 | -8·43±1·94 | 2·67 (-2·03 - 7·37) | 0.263 | -2·4±93·1 | -223·5±133·6 | 221·0 (-100·4 – 542·5) | 0.176 |
| 29-32w | -4·30±1·40 | -6·88±1·98 | 2·58 (-2·21 - 7·36) | 0.289 | -71·8±94·6 | -155·6±136·3 | 83·8 (-243·8 – 411·3) | 0.614 |
| 33-36w | -4·46±1·43 | -8·06±2·01 | 3·60 (-1·28 - 8·47) | 0.147 | -13·6±97·0 | 76·3±137·1 | -89·9 (-421·5 – 241·6) | 0.593 |
| 37-40w | -4·69±1·45 | -11·42±2·05 | 6·73 (1·79 - 11·68) | 0.008 | -91·3±98·2 | -191·2±137·5 | 99·9 (-233·8– 433·5) | 0.555 |
| 41-44w | -5·38±1·46 | -13·43±2·08 | 8·04 (3·03 - 13·05) | 0.002 | -8·6±100·7 | -88·3±151·0 | 79·7 (-278·5 – 438·0) | 0.661 |
| 45-48w | -6·01±1·49 | -15·17±2·11 | 9·16 (4·06 - 14·26) | <0.001 | -77·2±103·7 | -234·8±147·7 | 157·7 (-198·7 – 514·0) | 0.384 |
| 49-51w (or the end) | -7·64±1·53 | -17·51±2·51 | 9·86 (4·07 - 15·66) | <0.001 | -177·2±103·5 | -506·0±167·5 | 328·8 (-60·0 – 717·6) | 0.097 |

Table S4. 95%CI for Composite Z-score Analyses (Respiratory Function and Muscle Strength).

| | Time points | 5w | 9w | 13w | 17w | 21w | 24w | 26-27w | 29-31w | 33-35w | 37-39w | 41-43w | 45-47w | 48-50w<br>(or the end) |
|---|---|---|---|---|---|---|---|---|---|---|---|---|---|---|
| **%FVC** | **Difference‡(%)<br>(95%CI)<br>P value** | NA | NA | 2·8<br>(-6·1 - 11·7)<br>0.516 | NA | NA | 8·4<br>(-4·5 - 21·3)<br>0.190 | NA | NA | NA | 24·9<br>(3·1 - 46·7)<br>0.028 | NA | NA | 31·0<br>(-1·6 - 63·7)<br>0.061 |
| **MNS** | **Difference‡(points)<br>(95%CI)<br>P value** | -0·3<br>(-4·9 - 4·2)<br>0·886 | -0·7<br>(-5·2 - 3·9)<br>0·770 | -0·7<br>(-5·2 - 3·9)<br>0·774 | 0·2<br>(-4·3 - 4·7)<br>0·924 | 0·8<br>(-3·7 - 5·3)<br>0·724 | 3·6<br>(-0·9 - 8·2)<br>0·119 | NA | 2·0<br>(-2·6 - 6·6)<br>0·400 | 3·4<br>(-1·3 - 8·0)<br>0·158 | 3·1<br>(-1·6 - 7·8)<br>0·197 | 4·9<br>(0·1 - 9·8)<br>0·046 | 6·8<br>(1·8 - 11·8)<br>0·008 | 9·4<br>(3·9 - 14·9)<br><0·001 |
| **QMS: tongue pressure** | **Difference‡(kPa)<br>(95%CI)<br>P value** | -0·36<br>(-5·00 - 4·28)<br>0·879 | 0·56<br>(-4·09 - 5·20)<br>0·813 | 1·15<br>(-3·49 - 5·79)<br>0·625 | 0·44<br>(-4·20 - 5·08)<br>0·852 | 0·99<br>(-3·66 - 5·65)<br>0·673 | 1·74<br>(-3·02 - 6·51)<br>0·470 | NA | 1·22<br>(-3·67 - 6·11)<br>0·624 | 2·29<br>(-2·67 - 7·24)<br>0·363 | 3·61<br>(-1·41 - 8·64)<br>0·157 | -0·12<br>(-5·34 - 5·10)<br>0·964 | -4·67<br>(-9·97 - 0·63)<br>0·084 | -1·25<br>(-7·48 - 4·97)<br>0·691 |
| **QMS: elbow flexion (right)** | **Difference‡(N)<br>(95%CI)<br>P value** | -18·21<br>(-52·68 - 16·25)<br>0·298 | -9·57<br>(-44·04 - 24·89)<br>0·584 | -11·53<br>(-46·00 - 22·93)<br>0·510 | -18·32<br>(-52·78 - 16·15)<br>0·295 | -16·06<br>(-50·62 - 18·50)<br>0·360 | -28·70<br>(-63·66 - 6·25)<br>0·107 | NA | -11·34<br>(-47·07 - 24·38)<br>0·531 | 0·27<br>(-36·03 - 36·56)<br>0·988 | 13·79<br>(-22·86 - 50·44)<br>0·458 | 10·94<br>(-26·55 - 48·43)<br>0·565 | 12·22<br>(-26·16 - 50·60)<br>0·530 | 19·99<br>(-24·63 - 64·61)<br>0·377 |
| **QMS: elbow flexion (left)** | **Difference‡(N)<br>(95%CI)<br>P value** | -7·73<br>(-45·23 - 29·77)<br>0·684 | 18·38<br>(-19·12 - 55·88)<br>0·334 | 6·16<br>(-31·34 - 43·66)<br>0·746 | 12·43<br>(-25·07 - 49·93)<br>0·514 | 12·36<br>(-25·23 - 49·94)<br>0·517 | 17·49<br>(-20·41 - 55·39)<br>0·363 | NA | 40·57<br>(2·08 - 79·06)<br>0·039 | 28·86<br>(-9·87 - 67·58)<br>0·143 | 48·54<br>(9·86 - 87·22)<br>0·014 | 37·60<br>(-1·87 - 77·08)<br>0·062 | 49·13<br>(8·85 - 89·40)<br>0·017 | 35·49<br>(-10·05 - 81·03)<br>0·126 |
| **QMS: ankle dorsiflexion (right)** | **Difference‡(N)<br>(95%CI)<br>P value** | 27·32<br>(-37·92 - 92·56)<br>0·409 | 60·29<br>(-4·95 - 125·53)<br>0·070 | 69·29<br>(4·05 - 134·53)<br>0·038 | 41·98<br>(-23·26 - 107·22)<br>0·206 | 69·79<br>(4·37 - 135·21)<br>0·037 | 45·60<br>(-20·51 - 111·71)<br>0·175 | NA | 96·91<br>(29·64 - 164·17)<br>0·005 | 111·55<br>(43·77 - 179·34)<br>0·001 | 116·65<br>(47·41 - 185·88)<br>0·001 | 105·05<br>(34·74 - 175·37)<br>0·004 | 103·58<br>(32·93 - 174·24)<br>0·004 | 94·68<br>(9·62 - 179·74)<br>0·029 |
| **QMS: ankle dorsiflexion (left)** | **Difference‡(N)<br>(95%CI)<br>P value** | 7·59<br>(-38·20 - 53·38)<br>0·744 | 39·29<br>(-6·50 - 85·08)<br>0·092 | 22·56<br>(-23·23 - 68·36)<br>0·332 | 16·94<br>(-28·85 - 62·74)<br>0·466 | 21·54<br>(-24·48 - 67·55)<br>0·357 | 35·07<br>(-11·80 - 81·95)<br>0·141 | NA | 61·21<br>(12·82 - 109·59)<br>0·014 | 52·27<br>(3·43 - 101·11)<br>0·036 | 84·30<br>(33·78 - 134·81)<br>0·001 | 59·97<br>(8·53 - 111·41)<br>0·023 | 67·11<br>(14·58 - 119·64)<br>0·013 | 134·39<br>(63·35 - 205·42)<br><0·001 |
| **Body weight** | **Difference‡(kg)<br>(95%CI)<br>P value** | -0·38<br>(-4·70 - 3·93)<br>0·861 | -1·49<br>(-5·81 - 2·82)<br>0·495 | -2·08<br>(-6·40 - 2·23)<br>0·341 | -2·03<br>(-6·34 - 2·28)<br>0·354 | -1·55<br>(-5·87 - 2·76)<br>0·478 | -2·54<br>(-6·88 - 1·79)<br>0·248 | -1·57<br>(-5·95 - 2·80)<br>0·479 | -0·17<br>(-4·61 - 4·27)<br>0·941 | 0·42<br>(-4·10 - 4·95)<br>0·854 | 1·93<br>(-2·70 - 6·55)<br>0·412 | 2·11<br>(-2·61 - 6·84)<br>0·379 | 2·54<br>(-2·41 - 7·50)<br>0·313 | 3·08<br>(-2·36 - 8·52)<br>0·265 |
| **Serum creatinine** | **Difference‡(mg/dl)<br>(95%CI)<br>P value** | 0·019<br>(-0·048 - 0·086)<br>0·572 | 0·012<br>(-0·055 - 0·079)<br>0·718 | 0·005<br>(-0·062 - 0·073)<br>0·872 | -0·018<br>(-0·085 - 0·049)<br>0·601 | -0·004<br>(-0·072 - 0·063)<br>0·904 | -0·003<br>(-0·072 - 0·067)<br>0·940 | NA | -0·027<br>(-0·099 - 0·046)<br>0·466 | 0·005<br>(-0·069 - 0·079)<br>0·897 | 0·038<br>(-0·037 - 0·113)<br>0·318 | 0·091<br>(0·012 - 0·170)<br>0·024 | 0·105<br>(0·027 - 0·184)<br>0·009 | 0·052<br>(-0·041 - 0·145)<br>0·272 |

Table S5. 95%CI for Composite Z-score Analyses (ALSFRS-R Subdomains Score).

| ALSFRS-R subdomain score | Time points | 5w | 9w | 13w | 17w | 21w | 24w | 26-27w | 29-31w | 33-35w | 37-39w | 41-43w | 45-47w | 48-50w (or the end) |
|---|---|---|---|---|---|---|---|---|---|---|---|---|---|---|
| **Bulbar function** | **Difference‡ (points)** | 0·1 | -0·1 | -0·1 | -0·1 | 0·2 | 1·0 | 0·9 | 0·8 | 1·3 | 1·8 | 2·8 | 2·7 | 3·4 |
| | **(95%CI)** | (-1·2 to 1·3) | (-1·4 to 1·1) | (-1·3 to 1·1) | (-1·3 to 1·2) | (-1·1 to 1·4) | (-0·3 to 2·2) | (-0·4 to 2·2) | (-0·5 to 2·2) | (0·0 to 2·7) | (0·4 to 3·1) | (1·4 to 4·2) | (1·2 to 4·2) | (1·6 to 5·3) |
| | **P value** | 0·903 | 0·821 | 0·876 | 0·931 | 0·802 | 0·138 | 0·196 | 0·213 | 0·052 | 0·013 | <0·001 | <0·001 | <0·001 |
| **Motor function** | **Difference‡ (points)** | 0·3 | -0·1 | 0·0 | 0·1 | 0·5 | 1·1 | 2·2 | 2·2 | 2·5 | 5·0 | 5·3 | 6·5 | 7·0 |
| | **(95%CI)** | (-3·2 - 3·7) | (-3·6 - 3·3) | (-3·4 - 3·4) | (-3·3 - 3·5) | (-2·9 - 4·0) | (-2·4 - 4·6) | (-1·3 - 5·8) | (-1·4 - 5·7) | (-1·1 - 6·1) | (1·3 - 8·6) | (1·5 - 9·0) | (2·6 - 10·3) | (2·7 - 11·4) |
| | **P value** | 0·875 | 0·945 | 1·000 | 0·955 | 0·764 | 0·535 | 0·215 | 0·237 | 0·178 | 0·009 | 0·006 | 0·001 | 0·002 |
| **Respiratory function** | **Difference‡ (points)** | -0·1 | 0·1 | -0·5 | -0·6 | 0·2 | -0·2 | -0·3 | -0·4 | -0·3 | -0·1 | -0·2 | 0·1 | -0·2 |
| | **(95%CI)** | (-1·0 - 0·9) | (-0·9 - 1·0) | (-1·4 - 0·5) | (-1·6 - 0·4) | (-0·8 - 1·1) | (-1·2 - 0·8) | (-1·4 - 0·8) | (-1·5 - 0·7) | (-1·4 - 0·8) | (-1·2 - 1·0) | (-1·2 - 0·9) | (-1·0 - 1·2) | (-1·3 - 0·9) |
| | **P value** | 0·892 | 0·875 | 0·356 | 0·216 | 0·717 | 0·708 | 0·582 | 0·483 | 0·568 | 0·816 | 0·774 | 0·854 | 0·769 |

Table S6. 95%CI for Composite Z-score Analyses (Muscle Strength, Grip Strength, and Pinch Strength).

| | Time points | 5w | 9w | 13w | 17w | 21w | 24w | 29-31w | 33-35w | 37-39w | 41-43w | 45-47w | 48-50w (or the end) |
|---|---|---|---|---|---|---|---|---|---|---|---|---|---|
| **QMS: wrist extension (right)** | **Difference‡(N)** | 0·03 | -6·53 | -6·93 | -9·82 | -4·95 | 2·40 | -6·88 | 4·01 | 2·22 | 31·07 | 13·09 | 51·53 |
| | **(95%CI)** | (-42·31 - 42·37) | (-48·87 - 35·81) | (-49·27 - 35·41) | (-52·16 - 32·52) | (-47·41 - 37·50) | (-40·47 - 45·27) | (-50·43 - 36·68) | (-39·84 - 47·87) | (-42·02 - 46·46) | (-14·63 - 76·77) | (-33·44 - 59·62) | (-4·86 - 107·92) |
| | **P value** | 0·999 | 0·761 | 0·747 | 0·647 | 0·818 | 0·912 | 0·756 | 0·857 | 0·921 | 0·181 | 0·579 | 0·073 |
| **QMS: wrist extension (left)** | **Difference‡(N)** | 5·52 | 0·27 | -2·58 | 15·29 | 6·52 | 13·42 | 21·28 | 33·24 | 76·20 | 54·26 | 51·02 | 76·36 |
| | **(95%CI)** | (-27·44 - 38·49) | (-32·70 - 33·23) | (-35·54 - 30·39) | (-17·68 - 48·25) | (-26·67 - 39·71) | (-20·75 - 47·59) | (-14·11 - 56·67) | (-2·64 - 69·11) | (39·91 - 112·50) | (15·57 - 92·96) | (11·43 - 90·60) | (24·87 - 127·86) |
| | **P value** | 0·741 | 0·987 | 0·878 | 0·361 | 0·699 | 0·439 | 0·237 | 0·069 | <0·001 | 0·006 | 0·012 | 0·004 |
| **QMS: hip flexion (right)** | **Difference‡(N)** | 24·64 | 18·07 | 16·08 | 20·47 | 33·99 | 27·62 | 47·30 | 63·85 | 76·91 | 94·37 | 66·76 | 70·05 |
| | **(95%CI)** | (-29·38 - 78·65) | (-35·94 - 72·09) | (-37·93 - 70·10) | (-33·55 - 74·48) | (-20·22 - 88·19) | (-27·28 - 82·52) | (-8·72 - 103·33) | (7·28 - 120·43) | (19·83 - 133·98) | (35·02 - 153·73) | (5·99 - 127·53) | (10·67 - 129·43) |
| | **P value** | 0·369 | 0·510 | 0·557 | 0·455 | 0·217 | 0·322 | 0·097 | 0·027 | 0·009 | 0·002 | 0·032 | 0·021 |
| **QMS: hip flexion (left)** | **Difference‡(N)** | 18·46 | 10·84 | 35·52 | 10·73 | 25·70 | 15·08 | 62·24 | 69·92 | 88·39 | 97·08 | 52·56 | 78·56 |
| | **(95%CI)** | (-20·28 - 57·20) | (-27·90 - 49·58) | (-3·22 - 74·26) | (-28·01 - 49·47) | (-13·22 - 64·63) | (-24·58 - 54·74) | (21·17 - 103·30) | (28·02 - 111·83) | (46·15 - 130·63) | (53·15 - 141·02) | (8·09 - 97·04) | (20·40 - 136·73) |
| | **P value** | 0·348 | 0·581 | 0·072 | 0·585 | 0·194 | 0·454 | 0·003 | 0·001 | <0·001 | <0·001 | 0·021 | 0·008 |
| **QMS: grip (right)** | **Difference‡(kg)** | -0·82 | -2·75 | -2·28 | -2·25 | -1·94 | -2·61 | -2·89 | -4·73 | -4·47 | -1·88 | -1·49 | -2·79 |
| | **(95%CI)** | (-4·54 - 2·90) | (-6·47 - 0·97) | (-6·00 - 1·44) | (-5·98 - 1·47) | (-5·67 - 1·80) | (-6·40 - 1·18) | (-6·80 - 1·02) | (-8·71 – -0·75) | (-8·43 – -0·50) | (-5·80 - 2·05) | (-5·44 - 2·47) | (-7·04 - 1·46) |
| | **P value** | 0·663 | 0·146 | 0·228 | 0·233 | 0·307 | 0·176 | 0·146 | 0·020 | 0·027 | 0·347 | 0·460 | 0·197 |
| **QMS: grip (left)** | **Difference‡(kg)** | 1·65 | 2·84 | 3·93 | 3·75 | 5·28 | 2·41 | 4·89 | 3·32 | 3·11 | 4·99 | 6·41 | 6·20 |
| | **(95%CI)** | (-2·12 - 5·42) | (-0·93 - 6·61) | (0·16 - 7·71) | (-0·03 - 7·52) | (1·47 - 9·08) | (-1·53 - 6·36) | (0·71 - 9·08) | (-0·97 - 7·62) | (-1·21 - 7·42) | (0·56 - 9·42) | (1·84 - 10·98) | (0·10 - 12·30) |
| | **P value** | 0·388 | 0·139 | 0·041 | 0·052 | 0·007 | 0·229 | 0·022 | 0·128 | 0·157 | 0·028 | 0·006 | 0·046 |
| **QMS: pinch (right)** | **Difference‡(kg)** | -0·10 | -0·34 | -0·17 | -0·21 | -0·15 | -0·29 | -0·29 | 0·02 | 0·16 | -0·16 | -0·16 | -0·18 |
| | **(95%CI)** | (-0·92 - 0·71) | (-1·16 - 0·48) | (-0·98 - 0·65) | (-1·03 - 0·61) | (-0·97 - 0·67) | (-1·12 - 0·54) | (-1·14 - 0·55) | (-0·82 - 0·87) | (-0·69 - 1·02) | (-1·03 - 0·71) | (-1·05 - 0·72) | (-1·23 - 0·86) |
| | **P value** | 0·801 | 0·410 | 0·689 | 0·615 | 0·720 | 0·492 | 0·493 | 0·954 | 0·703 | 0·710 | 0·718 | 0·726 |
| **QMS: pinch (left)** | **Difference‡(kg)** | -0·11 | 0·20 | 0·66 | 0·42 | 0·43 | 0·72 | 0·83 | 1·11 | 1·13 | 1·12 | 1·70 | 1·48 |
| | **(95%CI)** | (-0·79 - 0·56) | (-0·47 - 0·87) | (-0·01 - 1·33) | (-0·25 - 1·10) | (-0·24 - 1·11) | (0·03 - 1·41) | (0·11 - 1·55) | (0·38 - 1·84) | (0·39 - 1·86) | (0·35 - 1·89) | (0·92 - 2·48) | (0·52 - 2·44) |
| | **P value** | 0·738 | 0·556 | 0·054 | 0·215 | 0·207 | 0·041 | 0·024 | 0·003 | 0·003 | 0·005 | <0·001 | 0·003 |

Table S7. 95%CI for Composite Z-score Analyses (ALSAQ-40 Scores).

| | Time points | 5w | 9w | 13w | 17w | 21w | 24w | 26-27w | 29-31w | 33-35w | 37-39w | 41-43w | 45-47w | 48-50w (or the end) |
|---|---|---|---|---|---|---|---|---|---|---|---|---|---|---|
| **ALSAQ-40 (Physical mobility)** | **Difference (points)** | 5·1 | 2·0 | 0·5 | -1·4 | 0·7 | -2·0 | -2·0 | -4·5 | -0·1 | -7·8 | -4·2 | -4·4 | -6·7 |
| | **(95%CI)** | (-1·7 - 11·8) | (-4·7 - 8·8) | (-6·3 - 7·2) | (-8·2 - 5·3) | (-6·1 - 7·5) | (-8·9 - 4·9) | (-9·1 - 5·1) | (-11·7 - 2·7) | (-7·3 - 7·2) | (-15·2 - -0·4) | (-12·0 - 3·5) | (-12·3 - 3·5) | (-15·7 - 2·3) |
| | **P value** | 0.140 | 0.554 | 0.895 | 0.678 | 0.842 | 0.565 | 0.577 | 0.215 | 0.984 | 0.040 | 0.283 | 0.274 | 0.143 |
| **ALSAQ-40 (ADL/independence)** | **Difference (points)** | 1·9 | 3·6 | 3·3 | -0·8 | -1·5 | -2·0 | -0·3 | -3·0 | -0·7 | -5·3 | -0·9 | -4·7 | -4·8 |
| | **(95%CI)** | (-5·2 - 9·1) | (-3·5 - 10·7) | (-3·8 - 10·4) | (-7·9 - 6·3) | (-8·6 - 5·7) | (-9·2 - 5·2) | (-7·7 - 7·0) | (-10·4 - 4·4) | (-8·2 - 6·8) | (-12·9 - 2·2) | (-8·8 - 6·9) | (-12·6 - 3·1) | (-14·1 - 4·4) |
| | **P value** | 0.590 | 0.315 | 0.357 | 0.826 | 0.683 | 0.587 | 0.933 | 0.427 | 0.850 | 0.165 | 0.811 | 0.238 | 0.306 |
| **ALSAQ-40 (Eating and drinking)** | **Difference (points)** | -1·5 | -0·4 | 0·4 | 0·3 | -0·4 | -0·7 | -0·4 | -1·8 | -1·2 | -2·3 | -0·8 | -0·8 | -0·9 |
| | **(95%CI)** | (-4·2 - 1·2) | (-3·1 - 2·3) | (-2·3 - 3·1) | (-2·3 - 3·0) | (-3·1 - 2·3) | (-3·4 - 2·0) | (-3·1 - 2·4) | (-4·6 - 1·0) | (-4·1 - 1·6) | (-5·2 - 0·5) | (-3·7 - 2·1) | (-3·7 - 2·2) | (-4·3 - 2·4) |
| | **P value** | 0.268 | 0.758 | 0.765 | 0.808 | 0.756 | 0.600 | 0.796 | 0.217 | 0.384 | 0.107 | 0.568 | 0.606 | 0.593 |
| **ALSAQ-40 (Communication)** | **Difference (points)** | 1·1 | -0·6 | 0·9 | -1·3 | -1·0 | -0·2 | -0·6 | -2·1 | 0·5 | -2·1 | -1·3 | -2·4 | 0·9 |
| | **(95%CI)** | (-3·0 - 5·2) | (-4·7 - 3·4) | (-3·2 - 5·0) | (-5·4 - 2·8) | (-5·1 - 3·1) | (-4·4 - 3·9) | (-4·9 - 3·7) | (-6·5 - 2·2) | (-3·9 - 4·9) | (-6·6 - 2·3) | (-6·0 - 3·4) | (-7·2 - 2·4) | (-5·4 - 7·3) |
| | **P value** | 0.596 | 0.758 | 0.656 | 0.524 | 0.618 | 0.912 | 0.779 | 0.337 | 0.830 | 0.350 | 0.583 | 0.328 | 0.773 |
| **ALSAQ-40 (Emotional functioning)** | **Difference (points)** | -0·3 | -3·6 | -3·6 | -5·6 | -3·4 | -5·8 | -4·7 | -4·1 | -4·2 | -6·6 | -5·2 | -4·5 | -7·2 |
| | **(95%CI)** | (-7·6 - 7·1) | (-10·9 - 3·8) | (-10·9 - 3·8) | (-13·0 - 1·8) | (-10·8 - 3·9) | (-13·3 - 1·6) | (-12·3 - 2·8) | (-11·7 - 3·6) | (-11·9 - 3·5) | (-14·4 - 1·2) | (-13·2 - 2·8) | (-12·6 - 3·7) | (-16·6 - 2·3) |
| | **P value** | 0.941 | 0.338 | 0.341 | 0.134 | 0.358 | 0.125 | 0.217 | 0.293 | 0.286 | 0.098 | 0.203 | 0.282 | 0.136 |

Table S8. Time to Death or Disease Progression Events (Entire Trial Period).

| Drugs | Participants | Weeks since the first dose | Details of disease progress |
|---|---|---|---|
| Ropinirole | No. | | |
| | 1 | 8·4 | Inability of independent ambulation |
| | 2 | 49·0 | Loss of unilateral upper limb function |
| | 3 | 53·4 | - |
| | 4 | 52·4 | - |
| | 5 | 55·3 | - |
| | 6 | 55·4 | - |
| | 7 | 50·3 | Respiratory support |
| | 8 | 23·0 | Inability of independent ambulation |
| | 9 | 23·0 | Inability of independent ambulation |
| | 10 | 46·3 | - |
| | 11 | 8·4 | Inability of independent ambulation |
| | 12 | 8·4 | Inability of independent ambulation |
| | 13 | 27·3 | - |
| Placebo | No. | | |
| | 1 | 8·4 | Inability of independent ambulation |
| | 2 | 37·4 | Inability of independent ambulation |
| | 3 | 4·3 | Loss of useful speech |
| | 4 | 26·0 | Inability of independent ambulation |
| | 5 | 22·4 | Death |
| | 6 | 41·4 | Inability of independent ambulation, Loss of useful speech |
| | 7 | 4·3 | Inability of independent ambulation |

Table S9. Effect of Ropinirole on the Rate of Disease Progression.

(A) The spreadsheet of death or disease progression events (entire trial period).

| **Event** | **Placebo (n=7)** | **Ropinirole (n=13)** | **Total** |
|---|---|---|---|
| Death | 1 | 0 | 1 |
| Disability of independent ambulation | 5 | 5 | 10 |
| Loss of upper limb function | 0 | 1 | 1 |
| Tracheotomy | 0 | 0 | 0 |
| Use of respirator | 0 | 1 | 1 |
| Use of tube feeding | 0 | 0 | 0 |
| Loss of useful speech | 2 | 0 | 2 |
| | | | 15 |
| **Number of subjects with events** | 7 (100%) | 7 (54%) | |

(B) The spreadsheet of Japanese ALS severity scale categories of participants at the end of 24 weeks and 48 weeks.

| Treatment group | ALS Severity grade at baseline | At the end of double-blind phase‡ | | | | | | At the end of open-label phase‡ | | | | | |
|---|---|---|---|---|---|---|---|---|---|---|---|---|---|
| | | 1 | 2 | 3 | 4 | 5 | Total | 1 | 2 | 3 | 4 | 5 | Total |
| Placebo | 1 | 0 | 2 | 0 | 1 | 0 | 3 | 0 | 0 | 0 | 2 | 1 | 3 |
| | 2 | 0 | 0 | 0 | 3 | 0 | 3 | 0 | 0 | 0 | 0 | 1 | 1 |
| | 4 | 0 | 0 | 0 | 1 | 0 | 1 | 0 | 0 | 0 | 1 | 0 | 1 |
| | **Total** | **0** | **2** | **0** | **5** | **0** | **7** | **0** | **0** | **0** | **3** | **2** | **5** |
| Ropinirole | 1 | 1 | 1 | 0 | 1 | 1 | **4** | 2 | 0 | 0 | 1 | 1 | **4** |
| | 2 | 1 | 3 | 0 | 4 | 0 | **8** | 1 | 2 | 0 | 4 | 0 | **7** |
| | 4 | 0 | 0 | 0 | 1 | 0 | **1** | 0 | 0 | 0 | 1 | 0 | **1** |
| | **Total** | **2** | **4** | **0** | **6** | **1** | **13** | **3** | **2** | **0** | **6** | **1** | **12** |

The Japan ALS severity classification promulgated by the Specified Disease Treatment Research Program for ALS of the Ministry of Health, Labor, and Welfare of Japan defines severity grades from 1 to 5, as follows:

1. Able to work or perform housework
2. Independent living but unable to work
3. Requiring assistance for eating, excretion or ambulation
4. Presence of respirator insufficiency, difficulty in coughing out sputum or dysphagia
5. Using a tracheostomy tube, tube feeding or tracheostomy positive pressure ventilation.

Table S10. The list of adverse effects in the previous phase 1 studies of Requip tablets in healthy individuals.
(Reference: Outline of application materials of Requip tablets (2006/Oct/20))

Abbreviations: AEs; adverse effects, BID; bis in die.

| Number of participants | Dosage | AEs |
|---|---|---|
| 9 | 0·4mg/single dosage | 1) Bad feeling (moderate), Cold sweat (moderate), Astasia (moderate)<br>2) Dizziness (mild)<br>3) Cold sweat (moderate), Dizziness (moderate), Astasia (moderate)<br>4) Cold sweat (moderate), Dizziness (moderate), Astasia (moderate)<br>5) Dizziness (mild)<br>All AEs are presumed to be derived from orthostatic hypotension. |
| 9 | 0·1, 0·2mg/single dosage | 1) Headache (mild) (0·1mg)<br>2) Headache (mild) (0·1mg)<br>3) Dizziness (mild) (0·1mg)<br>4) Drowsiness (mild) (0·2mg)<br>5) Cough, Drowsiness (mild) (0·2mg) |
| 8 | 0·2, 0·4, 0·6, 0·8mg/BID increased every 2 days | 1) Dizziness (mild) (0·2mg)<br>2) Dull headache, Dizziness (mild) (0·6mg) |
| 5 | 0·25mg/single dosage | None |

Table S11. Rates of Participant Disposition.

A. Percentage of discontinued cases from the time of main registration to the final observation at the 24th week of the double-blind period (analysis target: SAF)

| | Evaluation cases | Discontinued cases | Percentage of discontinued cases | | Difference of percentage of discontinued cases | |
|---|---|---|---|---|---|---|
| | N | N | % | 95%CI | % | 95%CI |
| Ropinirole | 13 | 1 | 7.7 | 0.2 - 36.0 | -6.6 | -36.3 - 23.1 |
| Placebo | 7 | 1 | 14.3 | 0.4 - 57.9 | - | - |

B. Percentage of discontinued cases from double-blind Day 1 to final evaluation of open-label extension period (analysis target: SAF)

| | Evaluation cases | Discontinued cases | Percentage of discontinued cases | | Difference of percentage of discontinued cases | |
|---|---|---|---|---|---|---|
| | N | N | % | 95%CI | % | 95%CI |
| Ropinirole | 13 | 6 | 46.2 | 19.2 - 74.9 | -39.6 | -77.1 - -2.1 |
| Placebo | 7 | 6 | 85.7 | 42.1 - 99.6 | - | - |
